# Adaptive nuclear GTP synthesis promotes glioblastoma treatment resistance and is a targetable vulnerability in patients

**DOI:** 10.1101/2025.11.11.25340023

**Authors:** Ningning Liang, Andrew J. Scott, Wajd N. Al-Holou, Jie Xu, Alexandra O’Brien, Angelica Lin, Vidhi Pareek, Zhou Sha, Zitong Zhao, John Z. Yang, Annabel Yang, Ayesha U. Kothari, Kari Wilder-Romans, Zhe Wu, Jiane Feng, Navyateja Korimerla, Ameer Elaimy, Sravya Palavalasa, Jiali Chen, Lu Wang, Li Zhang, Anthony C. Andren, Peter Sajjakulnukit, Bernard Marini, Jason A. Heth, Meredith A. Morgan, Theodore S. Lawrence, Stephen J. Benkovic, Deepak Nagrath, Sriram Chandrasekaran, Nathan R. Qi, Duxin Sun, Bo Wen, Manjunath P. Pai, Nathan Clarke, Krithika Suresh, Yoshie Umemura, Costas A. Lyssiotis, Weihua Zhou, Daniel R. Wahl

**Author notes:** Equal contribution (Ningning Liang, Andrew J. Scott, Wajd N. Al-Holou).

## Abstract

Glioblastoma (GBM), the most lethal of all brain cancers, resists therapy by rerouting metabolism and GTP signaling to promote DNA repair and radiotherapy (RT) resistance. How GBM modulates GTP levels after RT-induced DNA damage and whether this metabolic activity is therapeutically tractable alongside standard-of-care chemoradiation therapy remain unclear. Here, we identify rapid metabolic changes within hours of RT, including post-RT rewiring of guanylate synthesis driven by nuclear translocation of the rate-limiting guanylate synthesis enzyme IMPDH1. This re-localization and nuclear GTP accumulation depend on the DNA damage kinase DNA-PK. Targeting GTP synthesis with mycophenolate mofetil (MMF) slows DNA damage repair and extends survival of intracranial murine models treated with chemoradiation. In a phase 0 clinical trial, oral MMF administration achieved active intracranial drug concentrations and target engagement in recurrent GBM tumors. These findings implicate IMPDH as a metabolic target in GBM whose inhibition is feasible and could complement chemoradiation therapy.

**One Sentence Summary:** This work provides the first demonstration of radiation-induced, DNA-PK-dependent nuclear IMPDH1 localization and GTP synthesis as a pro-survival mechanism in GBM.

## INTRODUCTION

Glioblastoma (GBM) is the deadliest form of brain cancer, uniformly recurring after standard-of-care radiation therapy (RT) and temozolomide (TMZ) chemotherapy. This aggressive recurrence leads to dismal outcomes, due in part to GBM rewiring of metabolic activity. Indeed, alterations in numerous metabolic pathways including the TCA cycle (*1, 2*), nucleotide synthesis (*3–5*), and amino acid metabolism (*6, 7*) promote GBM phenotypes including stemness, growth, invasion, and immunosuppression.

Altered metabolism also facilitates GBM treatment resistance, and a variety of metabolic pathways are linked to glioma RT sensitivity (*8, 9*). Purines, especially GTP, are used by GBM cells to promote DNA repair and RT resistance via GTP-mediated signaling (*10, 11*). GBMs also rewire GTP production to resist TMZ, the standard-of-care chemotherapy conventionally administered alongside RT (*12*). In other cancers, GTP and GTP-synthesizing enzymes promote resistance to genotoxic chemotherapies (*13, 14*). These prior findings point to GTP production as a key mediator of resistance to DNA-damaging therapies in cancer.

While GTP promotes DNA repair and RT resistance, it is less clear whether RT modulates GTP levels, or if already-existing GTP pools are sufficiently radioprotective. DNA damage can activate ribonucleotide reductase, enabling the synthesis of deoxynucleotides needed as building material to repair broken DNA (*15*). In breast cancers, genotoxic chemotherapy can activate pyrimidine synthesis through an ERK-mediated phosphorylation of the rate-limiting enzyme in *de novo* pyrimidine synthesis (*16*). Genotoxic chemotherapy can also alter activity of inosine monophosphate dehydrogenase (IMPDH), the rate-limiting enzyme of *de novo* GTP biosynthesis. A recent study in triple-negative breast cancer has reported that endogenous DNA damage promotes translocation of the IMPDH2 isoform to the nucleus, where it may regulate nuclear NAD^+^ levels, PARP activity, and DNA repair (*13*). This chemotherapy-induced translocation of IMPDH2 does not appear to depend on the DNA damage response. How and whether nucleotides in general and GTP in particular respond to DNA damage in GBM is uncertain.

These biologic observations have begun to move towards practical application. Pharmacologic inhibitors of IMPDH, including the FDA-approved drug mycophenolate mofetil (MMF), can enhance the efficacy of RT and temozolomide in mouse models of glioma (*11, 12, 17*) and have activity in brain metastases models (*18*). Whether these drugs have similar efficacy in GBM patients is not known. Moreover, it is not known whether existing IMPDH inhibitors effectively cross the blood-brain barrier and exert metabolic effects intracranially, which is a critical obstacle in glioma therapeutic development.

In this study, we identify acute changes in GBM purine metabolism that occur within hours of RT. We find that biosynthesis of guanylates, but not adenylates or pyrimidines, acutely increases immediately following RT in cell line models and patient-derived spheroid and murine models of GBM. These RT-induced changes are dependent on the DNA damage response and are reversed by pharmacologic and genetic inhibition of the apical kinase DNA-PK. Mechanistically, we find that IMPDH1 localizes in the nucleus following RT in a DNA-PK-dependent fashion, where it promotes the nuclear synthesis of GTP. Inhibiting this biology with MMF in intracranial GBM models lowers tumor GTP/IMP ratios, slows the repair of DNA damage induced by combined RT/TMZ, and augments the efficacy of these standard-of-care treatments. We then performed a phase 0 clinical trial in patients with recurrent GBM, which revealed that oral treatment with MMF led to active drug concentrations and depleted GTP/IMP ratios in tumors. Together these findings reveal new biology about how glioma cells metabolically adapt to DNA damage to fuel DNA repair and how targeting these links may be feasible in patients.

## RESULTS

### Subhead 1: Radiation therapy increases purine levels in cell lines and mouse models of GBM

Metabolic regulation of DNA repair occurs through numerous canonical and non-canonical mechanisms (*8*). How DNA damage alters metabolism is less understood, although we have previously correlated purine abundance broadly with RT resistance in immortalized cell lines (*11*). Using this information, we investigated the open questions of which metabolites change, and their magnitudes and pathway activities, post-RT as a starting point to glean novel mechanistic information. To identify metabolites that acutely change in response to RT-induced DNA damage, we quantified specific nucleotides and their precursors in our dataset profiling metabolomes of 4 cell lines under control conditions or 2 h post-RT. Increases in guanylate abundance after RT were observed across all 4 cell lines tested (Fig. 1A). These increases were both in deoxy- and non-deoxy guanylates, which suggested regulation separately from the known role of ribonucleotide reductase in the DNA damage response (*15*). RT-induced changes in other nucleotide species (adenylates, cytidylates, thymidylates, and uridylates) were more heterogeneous with varied increases and decreases in various cell lines (fig. S1A-S1D).

**Fig. 1.**
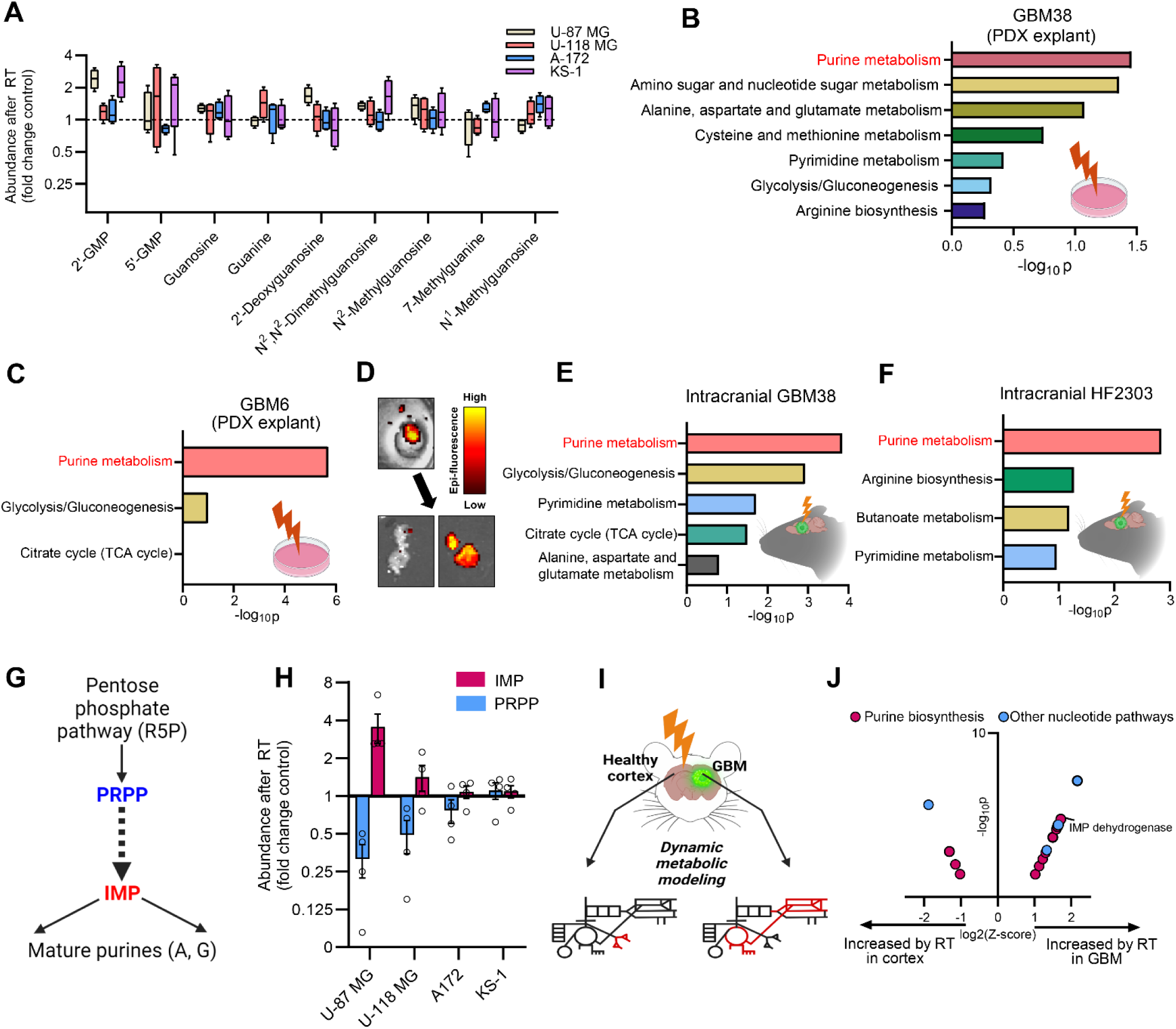
RT modulates GBM purine metabolites. (**A**) RT-sensitive (U-118 MG and KS-1) and RT-resistant (U-87 MG and A-172) GBM cell lines were irradiated (0 or 8 Gy), followed by targeted metabolomics of samples collected 2 hours after RT. n=4 biologically independent samples per group. (**B**, **C**) Metabolite pathway enrichment using differential metabolites 2 h after RT (8 Gy) in GBM38 cells (**B**) and GBM06 cells (**C**) explanted from PDX tissue. (**D**) Dissection of GFP-positive GBM tissue from GFP-negative brain tissue. (**E**, **F**) Metabolite pathway enrichment using differential metabolites after RT in orthotopic GBM38 (**E**) and orthotopic HF2303 (**F**) tumors. (**G**) Biosynthesis of *de novo* purines: pentose phosphate pathway (R5P)-derived PRPP undergoes a series of biosynthesis reactions to yield IMP, which is then converted into mature adenylates or guanylates. PRPP, phosphoribosyl pyrophosphate; IMP, inosine 5′-monophosphate. (**H**) Abundance of IMP and PRPP after RT in GBM cell lines from the dataset in (**A**). (**I**) Cortex and GBM tissues from intracranial GBM38-bearing mice were collected for metabolomics at multiple timepoints post-RT to predict differentially altered pathway activity via dynamic flux analysis (DFA). (**J**) Biochemical reactions predicted to increase after RT in mouse cortex and intracranial GBM38 tissue were determined by DFA using metabolite abundance measurements from tissues collected from RT-treated mice at multiple timepoints. Each datapoint represents a different enzymatic reaction.

To determine if these findings extended beyond conventional immortalized cell line models, we separately generated *in vitro* explants using GBM38 and GBM06 patient-derived xenograft (PDX) models from the Mayo Clinic Brain Tumor PDX National Resource (*19*) and measured their metabolite levels 2 h after RT. Metabolite set enrichment analysis identified purine metabolism as the most altered metabolic pathway in both models (Fig. 1B, Fig. 1C). These changes appeared to again be driven by guanylate increases, though some adenylates also increased (fig. S1E, fig. S1G). Pyrimidine changes varied between models, with several species increasing in GBM38 (fig. S1F) and minimal changes in GBM06 (fig. S1H). Analyses of explants 24 h after RT indicated purine metabolism was no longer the most altered metabolic pathway (fig. S1I, fig. S1J), suggesting that altered purine metabolism is an acute change following RT.

We then tested whether these RT-induced metabolite changes occurred *in vivo*. We used the patient-derived models GBM38 and HF2303 (*20*) grown orthotopically and which expressed luciferase (for intracranial detection in live mice) and GFP (for separation from cortex during tissue harvests). When tumor establishment was confirmed via bioluminescence imaging, mice were treated with whole-brain RT, and 2-4 h later tumor tissue was separated from uninvolved cortex under GFP guidance (Fig. 1D). Purine metabolism was again the most altered metabolic pathway in both GBM models acutely after RT (Fig. 1E, Fig. 1F), which was driven both by altered adenylate and guanylate metabolism (fig. S2A-S2B). While pyrimidine metabolism also changed following RT in GBM, those changes were less significant (Fig. 1E-F, fig. S2C-S2D). Mixed metabolic responses were observed in the non-tumor cortex, with some purine and pyrimidine metabolites increasing after RT and others decreasing (fig. S2E-S2J).

Changes in metabolite levels do not reliably predict changes in metabolic activity, as an increase in a metabolite’s abundance could be due to either its increased formation or its decreased consumption. To begin to understand how nucleotide metabolic activity changed in GBM models following RT, we separately analyzed an upstream metabolite in the purine synthesis pathway (phosphoribosyl pyrophosphate, PRPP) and a downstream metabolite (inosine monophosphate, IMP, Fig. 1G). There appeared to be a reciprocal relationship between an increase in IMP levels and a decrease in PRPP levels in RT-resistant cell lines defined previously (*11*), which would be consistent with increased purine synthesis following RT (Fig. 1H). We then employed a more sophisticated dynamic flux activity (DFA) model using changes in the 150+ metabolites measured in cortex and tumor from orthotopic GBM38-bearing mice (Fig. 1I) (*21–23*). DFA modeling predicted increased activity in 18 steps in purine synthesis in GBM following RT, with the greatest synthesis in IMPDH, the rate-limiting step in guanylate synthesis (Fig. 1J). Predicted upregulation of purine synthesis in non-tumor contralateral cortex was more modest.

### Subhead 2: Radiation therapy increases GBM de novo guanylate synthesis

To directly interrogate nucleotide synthesis following RT in GBM, we employed stable isotope tracing, in which “heavy” but non-radioactive metabolic isotopes are introduced into a biologic system and followed into their downstream fates using mass spectrometry (*24*). Pathways were traced with amide-labeled ^15^N-glutamine, as the amide nitrogen of glutamine is used to synthesize both purines and pyrimidines (Fig. 2A, fig. S3A). During two reactions of *de novo* IMP synthesis, ^15^N from labeled glutamine can be directly incorporated into the purine ring, yielding m+1 and m+2 isotopologues that are 1-2 mass units heavier than unlabeled IMP. While no additional nitrogen atoms are added in the reactions converting IMP to AMP, one additional ^15^N atom from glutamine can be incorporated while IMP is converted to GMP, yielding m+3 isotopologues. If metabolites are not fully tracer-saturated and if *de novo* GMP synthesis activity is greater than *de novo* IMP synthesis, then m+1 labeling of GMP (from ^15^N addition to unlabeled IMP) may exceed its m+3 labeling. A-172 glioma cells were incubated with ^15^N-glutamine and processed 4 hours after RT for LC-MS analysis, which revealed increased ^15^N labeling in GTP after RT (Fig. 2B). Labeled ATP percentages were essentially unchanged (Fig. 2C). In GBM38 explants, we similarly observed increased post-RT ^15^N labeling of GTP (Fig. 2D), but not ATP (Fig. 2E). We also examined ^15^N-glutamine-derived labeling of pyrimidine species following RT but did not observe an RT-induced label increase (fig. S3B-S3E).

**Fig. 2.**
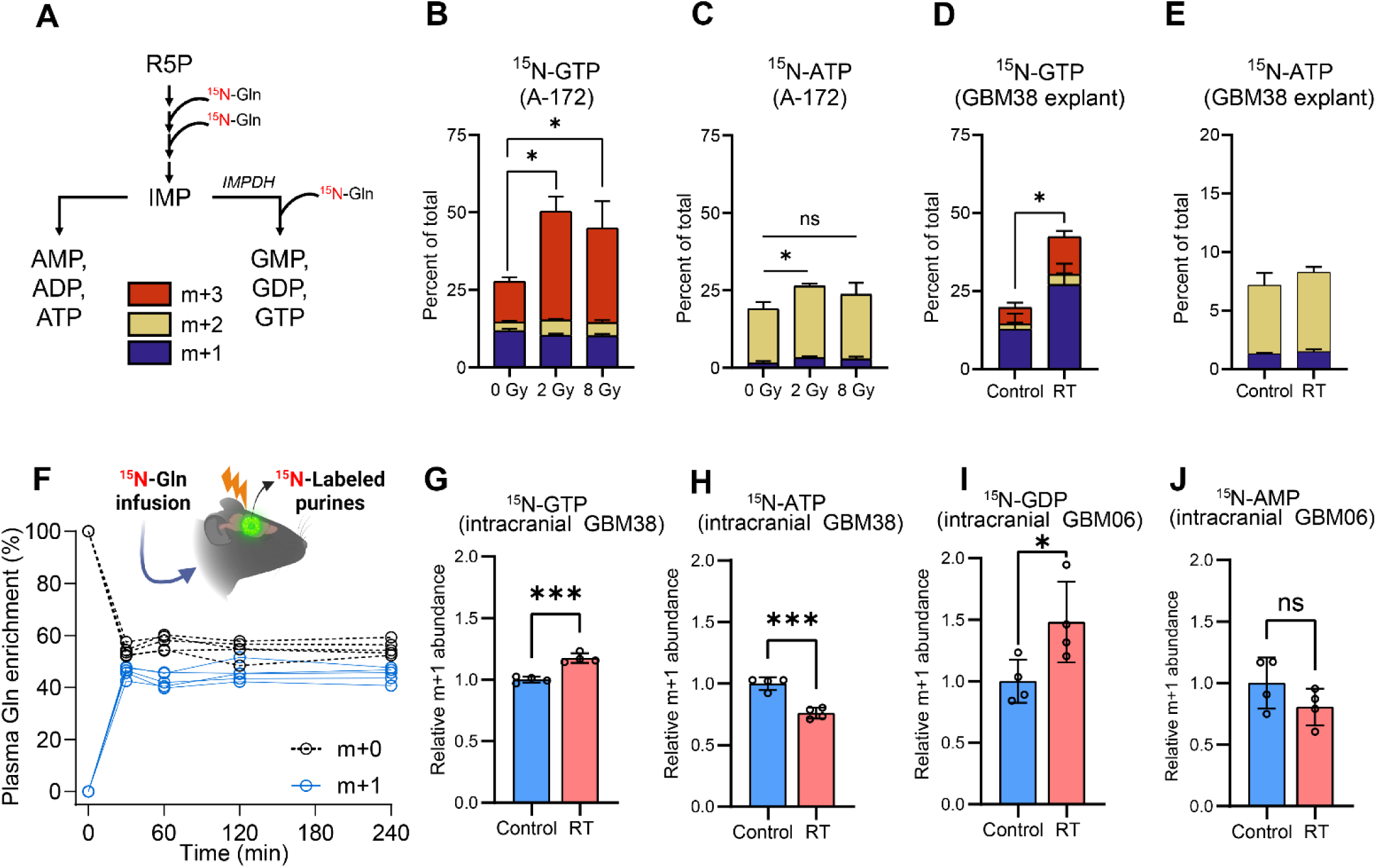
RT increases IMPDH-mediated guanylate synthesis in GBM. (**A**) Potential nitrogen labeling patterns of *de novo* purine intermediates from ^15^N-amide-glutamine. Prior to IMP, 2 glutamine nitrogen-incorporating reactions occur. One additional glutamine nitrogen may be added during IMPDH-mediated guanylate synthesis from IMP but not during adenylate synthesis from IMP. (**B**, **C**) Mass isotopologue distribution of ^15^N-glutamine-derived GTP (**B**) and ATP (**C**) in A-172 cells treated with indicated RT doses. Data are shown as mean ± SD from 4-5 biologically independent samples. (**D**, **E**) Mass isotopologue distribution of ^15^N-glutamine-derived GTP (**D**) and ATP (**E**) in GBM38 explants treated with 0 or 8 Gy RT. Data are shown as mean ± SD from 5 biologically independent samples. (**F**) Plasma levels of unlabeled (m+0) and labeled (m+1) glutamine in conscious, free-moving, undisturbed mice during intravenous infusions of ^15^N-glutamine. (**G**, **H**) Abundance of m+1 GTP (**G**) and ATP (**H**) in orthotopic GBM38 tumors harvested from mice treated with cranial RT (0 or 8 Gy) and immediately infused with ^15^N-glutamine for 4 h. Data are shown as mean ± SD from 4 biologically independent samples per group. (**I**, **J**) Abundance of m+1 GDP (**I**) and AMP (**J**) in orthotopic GBM06 tumors harvested from mice treated with cranial RT (0 or 8 Gy) and immediately infused with ^15^N-glutamine for 4 h. n=4 biological replicates per group. Data are shown as mean ± SD. *p*-values between groups were determined by two-tailed unpaired Student t-tests. * *p* < 0.05; ** *p* < 0.01; *** *p* < 0.001.

To determine if these metabolic events occur in GBMs within the native intracranial environment, we performed stable isotope infusion experiments using ^15^N-glutamine in unrestrained, conscious mice. Prior to infusions, mice underwent surgical procedures in which an intravenous catheter was placed for tracer administration, and an arterial catheter was placed for isolation of blood samples at multiple timepoints to determine plasma tracer enrichment (*25*). Using this approach, we determined our infusion parameters led to an arterial steady state in which approximately 40% of plasma glutamine contained ^15^N by 30 minutes after start of infusion and was consistent for the rest of the infusion duration (Fig. 2F). We then performed experiments in which intracranial GBM-bearing mice were treated with whole-brain RT immediately followed by infusion of ^15^N-glutamine. Consistent with *in vitro* studies, ^15^N-labeled GTP increased in GBM38 tumors following RT (Fig. 2G), whereas ^15^N-ATP modestly decreased (Fig. 2H). A similar RT-induced increase in ^15^N-guanylate labeling was observed in a second PDX model (GBM06, Fig. 2I). As in the GBM38 model, RT also decreased ^15^N-labeled adenylates in GBM06 tumors (Fig. 2J). RT did not increase ^15^N labeling of pyrimidines in either GBM38 or GBM06 intracranial tumors (fig. S3F-S3J).

Glutamine nitrogen can be used to synthesize purines *de novo* (Fig. 2A), but through the action of GMP synthase can label guanylates derived from both *de novo* and salvaged IMP. Thus, m+1 and m+2 guanylate isotopologues might originate from either pathway to fuel glioma metabolism (*25, 26*). To directly assess purine salvage after RT, stable isotope tracing experiments were performed using 2,8-^2^H_2_-hypoxanthine or 8-^13^C-guanine, both of which are substrates for the promiscuous salvage enzyme hypoxanthine-guanine phosphoribosyl transferase (HGPRT) with their co-substrate PRPP (fig. S3K, S3L). When 2,8-^2^H_2_-hypoxanthine and PRPP are substrates, both deuterium atoms label IMP and are carried into AMP (m+2), while one deuterium atom is lost during the IMPDH reaction driving synthesis of GMP (m+1). Alternatively, when 8-^13^C-guanine and PRPP are substrates, the labeled carbon remains within the intact purine ring and labels mature guanylates m+1. Neither hypoxanthine-nor guanine-derived labeling of purines increased following RT in GBM PDX explants (fig. S3M-O, fig. S3P). Together, these observations suggest that RT acutely increases the synthesis of guanine-containing purines, specifically via *de novo* pathways, in GBM *in vitro* and *in vivo*.

### Subhead 3: DNA damage signaling is responsible for increased guanylate synthesis after RT

We reasoned that RT-induced guanylate synthesis could either be (1) due to an active signaling event or (2) a compensatory upregulation of nucleotide production due to DNA damage. To determine which of these explanations accounted for upregulated guanylate synthesis, we inhibited key DNA damage-sensing kinases involved in the DNA damage response: DNA-dependent protein kinase (DNA-PK) using M3814, ataxia-telangiectasia mutated (*27*) using Ku60019, and ATM- and Rad3-Related (*27*) using AZD6738 (*28*). Activation of these proteins after RT and their inhibition after drug treatments were confirmed by immunoblots assessing protein phosphorylation (Fig. 3A). In GBM38 explants, pharmacologic inhibition of the catalytic subunit of DNA-PK (DNA-PKcs) stopped the increased synthesis of guanylates following RT (Fig. 3B) but had minimal effect on adenylate synthesis (Fig. 3C). Pharmacologic inhibition of ATM and ATR had less dramatic effects on RT-induced purine synthesis (fig. S4A-S4B). We also tested the HF2303 neurosphere model of GBM, where pharmacologic inhibition of DNA-PKcs with M3814 also blocked the RT-induced increase in guanylate synthesis (Fig. 3D). Changes in adenylate labeling in HF2303 cells were again modest (fig. S4C), though M3814 treatment appeared to slightly decrease adenylate labeling following RT treatment.

**Fig. 3.**
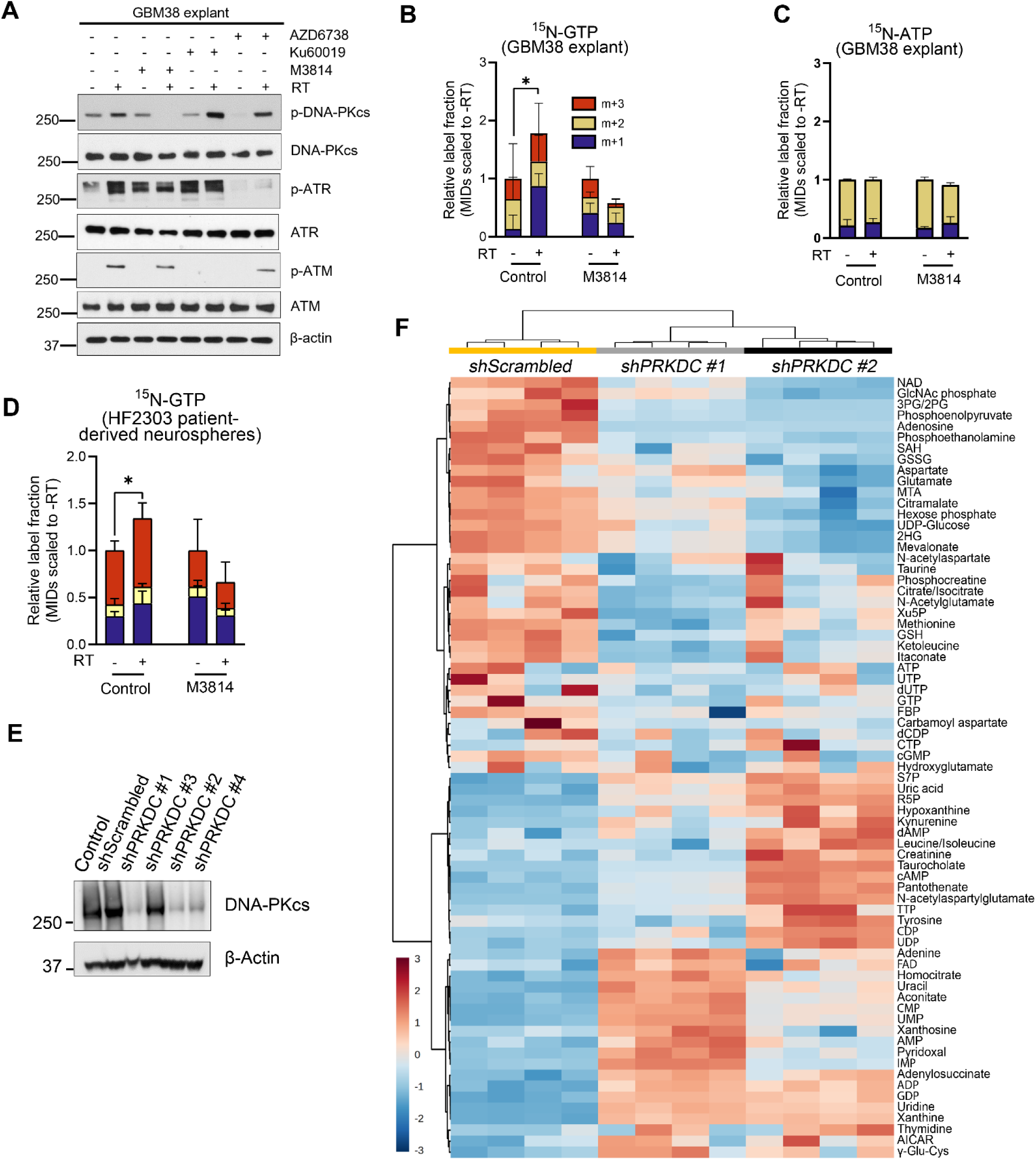
DNA damage signaling is responsible for increased IMPDH-derived guanylate synthesis after RT. (**A**) GBM38 explant cells treated with vehicle, M3814 (1 µM), KU60019 (2 µM), or AZD6738 (0.5 µM) 3 hours prior to RT (0 or 8 Gy) and harvested 1 h post-RT were probed for the indicated target proteins. (**B**, **C**) Mass isotopologue distributions of GTP (**B**) and ATP (**C**) in ^15^N-glutamine-labeled GBM38 explants treated with RT (0 or 8 Gy) and/or M3814 (0 or 1 µM). Data are shown as mean ± SD from 3-4 biologically independent samples per group. (**D**) Mass isotopologue distribution of GTP in ^15^N-glutamine-labeled HF2303 neurospheres treated with RT (0 or 8 Gy) and M3814 (0 or 1 µM). Data are shown as mean ± SD from 3-4 biologically independent samples per group. (**E**) Immunoblot for DNA-PKcs for GBM38 cells stably transduced with either non-targeting, scrambled shRNA (shScrambled) or shRNAs against *PRKDC* (shPRKDC) to determine extents of knockdown. Lines expressing shPRKDC constructs #1 and #2 were selected for further experimentation. (**F**) Heatmap of metabolite levels after RT in orthotopic GBM38 tumors proficient or deficient in DNA-PKcs. Values represent abundance ratios in RT-treated tumors over corresponding unirradiated groups of the same genotype. *p*-values between groups were determined by two-tailed unpaired Student t-tests. *, *p* < 0.05; **, *p* < 0.01; ***, *p* < 0.001.

We then sought to probe the role of DNA-PK signaling in RT-induced guanylate synthesis in GBM *in vivo*. We genetically suppressed DNA-PK in GBM38 explants using shRNAs targeting its encoding gene, *PRKDC* (Fig. 3E), and used these lines to establish intracranial tumors. Irradiated control (non-targeting shRNA, shScrambled) tumors exhibited altered abundance of numerous metabolites, including guanine-containing purines, compared to unirradiated control tumors (Fig. 3F). These RT-induced metabolic changes were largely inverted in tumors with silenced DNA-PKcs (Fig. 3F). To glean whether these changes were related to a DNA-PK link between RT and GTP synthesis, we examined metabolites upstream (ribose 5-phosphate, AICAR, IMP) and downstream (GTP) in guanylate synthesis. While upstream metabolites decreased following RT in control tumors (fig. S4D-S4E), this decrease was not observed in tumors with silenced DNA-PKcs. By contrast, there was a trend towards increased GTP levels following RT in control tumors that were absent in those with silenced DNA-PKcs (fig. S4G). These observations of upstream metabolite depletion and downstream metabolite accumulation are consistent with substrate consumption for increased GTP production requiring DNA-PK. Similar to prior experiments, ATP levels were not substantially impacted by RT in control tumors or DNA-PK-deficient tumors (fig. S4H). These results suggested a DNA-PKcs-dependent RT-induced production of guanylates in intracranial GBM tumors.

### Subhead 4: DNA-dependent protein kinase is required for nuclear IMPDH1 localization and catalysis after RT

Metabolic adaptation to stimuli can be controlled through several mechanisms. Because DNA-PKcs has kinase activity, we investigated whether RT caused a direct phosphorylation of the enzymes used to synthesize purines. Phosphoproteomic analysis of irradiated patient-derived GBM cells showed minimal changes in the phosphorylation states of purine-synthesizing enzymes (*10*) (fig. S5A). To further investigate this possibility, we used Phos-Tag gels to determine if RT altered phosphorylation-dependent SDS-PAGE mobility of enzymes with or without DNA-PK inhibition (*29*). Consistent with our phosphoproteomic studies, we did not observe substantial RT-induced phosphorylation changes or levels of enzymes mediating purine synthesis (fig. S5B).

To reconcile this lack of substantial change in metabolic enzyme levels/phosphorylation after RT with the altered metabolic activity observed in preceding experiments, we then investigated the spatial localization and kinetics of these enzymes. Purinosomes are dynamic multi-enzyme complexes that reversibly assemble and disassemble in the cytoplasm in response to purine levels and metabolic demands, enhancing *de novo* purine biosynthesis by channeling intermediates and restricting intermediate diffusion (*30, 31*). Proximity ligation assays (*32*) were performed to quantify colocalization of a pair of enzymes found in the purinosome, in which the enzymes’ close spatial proximity is identified by a PLA probe that generates fluorescent puncta when they are <40-60 nm apart as evidence for an assembled metabolon (*33*). When colocalization of phosphoribosylformylglycinamidine synthase (PFAS) and adenylosuccinate lyase (ADSL) was compared between irradiated and unirradiated cells fixed 4 h post-RT (0 or 8 Gy), no differences in puncta-positive proportions of either GBM38 or A-172 cells were observed (fig. S5C-S5D). However, complete purinosome assemblies require at least 6 different core purine biosynthesis enzymes in proximity and could be disrupted without impacting PFAS-ADSL proximity at the timepoint tested. Therefore, because PFAS-ADSL colocalization is a necessary but not sufficient condition for functional purinosomes, it remained possible that purinosome channeling could still be responsive to RT. Therefore, we also used isotopologue distributions from ^13^C_3_,^15^N-serine labeling of metabolites to estimate channeled purine biosynthesis output (*30, 34*). This analysis demonstrated significant decreases in purines generated via channeling in RT-treated cells, with less channeled synthesis of guanylates after RT in GBM38 cells and less channeled synthesis of both adenylates and guanylates after RT in A-172 cells (fig. S5E-S5F). The observation that guanylate synthesis channeling decreased after RT in both models implicated a shift specifically in the activity of IMPDH.

Data so far showing increased net *de novo* guanylate synthesis after RT and decreased cytoplasmic (mitochondria-adjacent) purinosome-mediated guanylate synthesis after RT suggested a subcellular compartment change for IMPDH activity. Because purinosome-channeled guanylate synthesis in the cytoplasm decreased and because RT-induced guanylate production presumably serves to facilitate the repair of DNA damage in the nucleus, we hypothesized that RT increases activity of nuclear IMPDH specifically. In GBM PDX explants and immortalized cell lines, IMPDH1 and IMPDH2 predominantly localized to the cytosol (Fig. 4A-4B). However, IMPDH1 abundance in the nucleus increased following RT that was blocked when cells were co-treated with M3814. While IMPDH1 and IMPDH2 are rate-limiting steps in guanylate synthesis, their enzymatic product xanthosine monophosphate must be further metabolized by GMP synthase (GMPS) to generate guanylates. Therefore, we also probed lysates for GMPS and found that GMPS was localized to both the cytosol and nucleus in both models (Fig. 4A-4B), suggesting the concept of post-RT nuclear IMPDH1 promoting nuclear guanylate formation is feasible. This RT-induced nuclear translocation also occurred in intracranial GBM PDX tumors and was similarly impaired when DNA-PKcs was knocked down (Fig. 4C-4D).

**Fig. 4.**
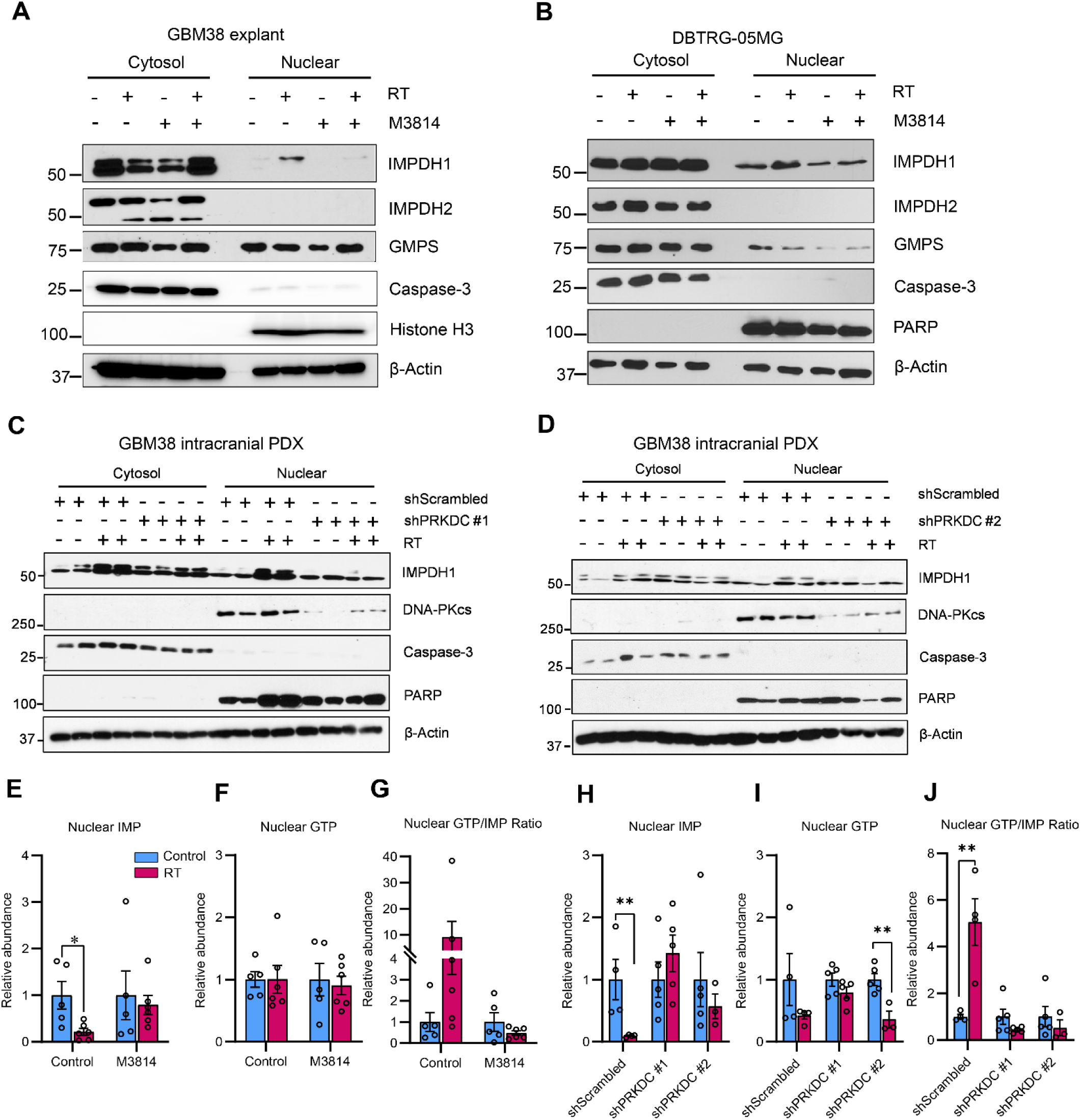
DNA-PK is required for nuclear IMPDH1 localization and catalysis after RT. (**A**, **B**) Cytosolic and nuclear fractions were assessed by immunoblot for the indicated proteins from GBM38 explants (**A**) or DBTRG-05MG (**B**) cells treated with RT (0 or 8 Gy) and/or M3814 (0 or 1 µM). (**C**, **D**) Intracranial GBM38 tumors stably transduced with control hairpin (shScrambled) vs. shPRKDC #1 (**C**) or shPRKDC #2 (**D**) were assessed for cytosolic and nuclear proteins by immunoblot after 0 or 8 Gy RT. (**E**-**G**) Nuclear fractions of GBM38 explants treated with M3814 (0 or 1 µM) or RT (0 or 8 Gy) were assessed for IMP (**E**) and GTP (**F**) by LC-MS and used to determine GTP/IMP ratios (**G**). Data were normalized to 0 Gy-treated vehicle/drug-only groups. n=5-6 biologically independent samples. Data are shown as mean ± SEM. (**H**-**J**) Nuclear fractions of intracranial GBM38-shScrambled or GBM38-shPRKDC tumors treated with RT (0 or 8 Gy) were assessed for IMP (**H**) and GTP (**I**) by LC-MS and used to determine GTP/IMP ratios (**J**). Data were normalized to 0 Gy-treated shScrambled tumors. Data are shown as mean ± SEM of n=3-5 biologically independent samples. *p*-values between groups were determined by two-tailed unpaired Student t-tests. *, *p* < 0.05; **, *p* < 0.01.

Our model suggested that RT-induced guanylate production (Fig. 1-3) might occur in the nucleus. Due to the challenges of compartment-specific isotope tracing, especially *in vivo*, we examined a metabolite upstream of IMPDH (IMP) and one downstream (GTP) as in Fig. 1G. RT acutely increased the ratio of downstream GTP to upstream IMP in the nucleus approximately 10-fold in control GBM38 PDX explants, consistent with increased nuclear guanylate synthesis activity.

This RT-induced change was reversed with pharmacologic DNA-PKcs inhibition (Fig. 4E-G), while the cytosolic GTP/IMP ratio was unaffected (fig. S5G-S5I). Similar observations were made *in vivo*, in which the nuclear GTP/IMP ratio increased approximately 5-fold in irradiated intracranial GBM tumors but not in DNA-PKcs-ablated tumors (Fig. 4H-J). Consistent with *in vitro* findings, the cytosolic GTP/IMP ratio was unaffected, although interestingly *in vivo* both IMP and GTP exhibited reduced cytosolic abundance (fig. S5J-S5L) that might indicate rapid extranuclear purine pathway activity attempting to compensate for IMPDH1 localization failure. These changes are consistent with RT acutely altering nuclear purine production in a DNA-PKcs-dependent fashion.

### Subhead 5: Targeting IMPDH with MMF can augment chemoradiation efficacy in GBM intracranial models

We then sought to target this biology to improve GBM therapy. We have previously found that guanylates promote DNA repair and genotoxic resistance through a unique signaling mechanism (*10, 11*). We therefore attempted to pharmacologically block GBM guanylate synthesis to slow DNA repair and improve responses to standard-of-care genotoxic radiation with temozolomide (RT/TMZ). Mycophenolate mofetil (MMF) is a prodrug of mycophenolic acid (MPA), which inhibits GTP synthesis by blocking IMPDH (*35*). We have previously found that MMF cooperates with RT in an orthotopic GBM model (*11*), and others have found similar benefits in combination with TMZ in similar models (*12*). The extent to which MMF treatment causes intracranial active drug accumulation in mouse models or human GBM patients is unknown. Similarly, we do not know how MMF treatment affects DNA repair in orthotopic GBM models or survival in combination with standard-of-care combined RT/TMZ.

To begin to address these questions, we established a liquid chromatography-mass spectrometry method to quantify MPA levels in tissue as well as metabolites upstream (IMP) and downstream (GTP) of the IMPDH reaction. We used three intracranial models including the human GBM38 PDX model (in immunodeficient Rag1 KO mouse hosts) (*19*). Because MMF is an immunosuppressive agent and could affect anti-tumor immunity, we also included syngeneic GL26 and TRP models (in immunocompetent C57/Bl6 mouse hosts) (*36, 37*). We treated tumor-bearing mice with oral MMF (150 mg/kg, once daily for 4 days) and 2-4 hours later harvested GFP-positive tumor and contralateral GFP-negative cortex for mass spectrometry analysis (fig. S6A). Oral MMF treatment yielded concentrations between 1.5-2.2 µM in cortical tissue and 14-21 µM in GBM tissue (Fig. 5A-5C)—levels previously shown to deplete purines in other models (*11, 38*). To assess intracranial target engagement of MMF, we also quantified the ratio of downstream GTP to upstream IMP, which is expected to decrease with IMPDH inhibition. When Rag1 KO mice with orthotopic GBM38 tumors were treated with MMF, GTP/IMP ratios decreased both in cortex and GBM tissue (Fig. 5D). When C57/BL6 mice bearing intracranial tumors were administered MMF, tumor GTP/IMP ratios trended downward in TRP tumors (fig. S6B) and appeared to decrease in GL26 (fig. S6C), although our analysis is limited by low sample number in this model (fig. S6B).

**Fig. 5.**
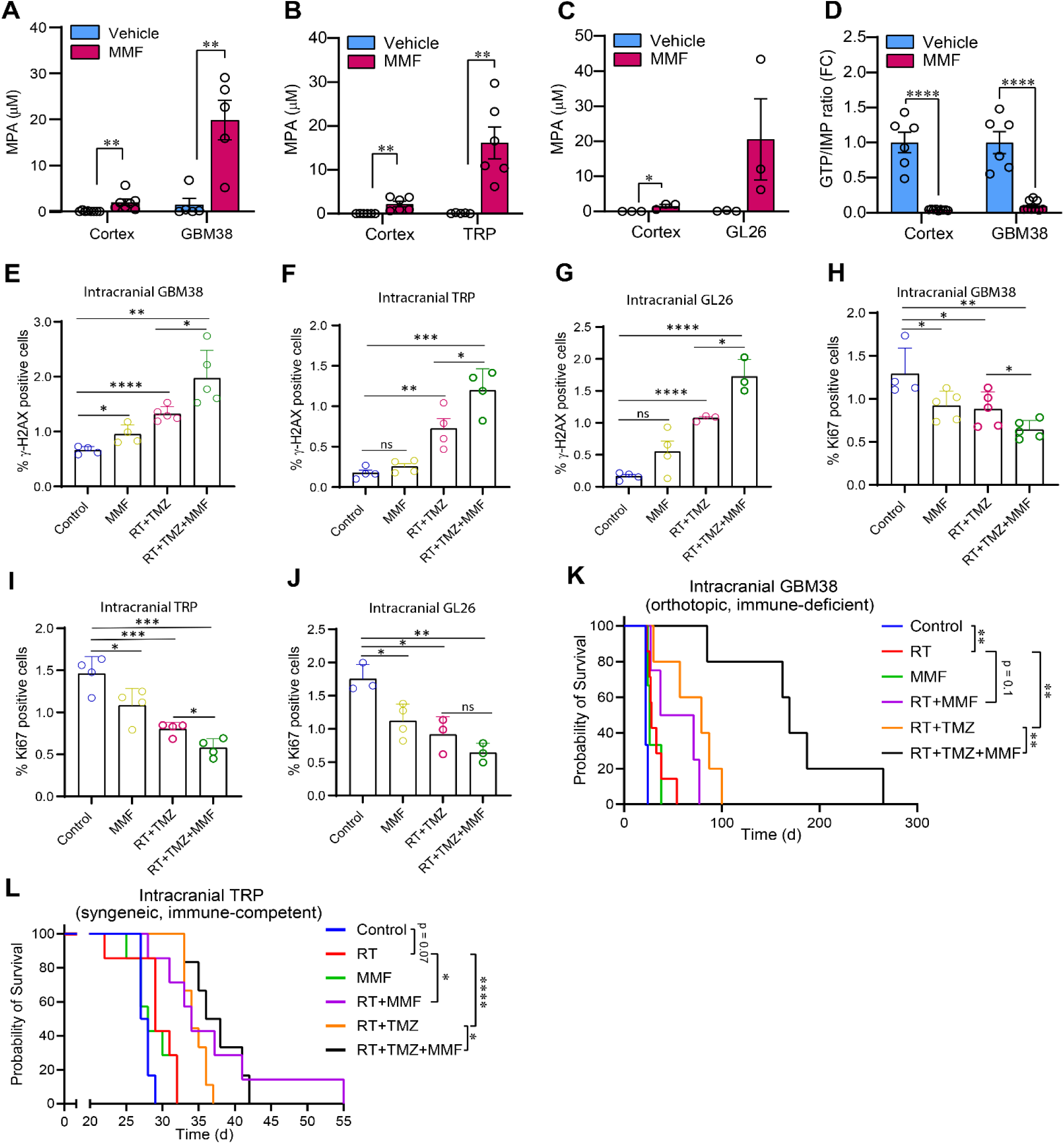
Targeting IMPDH activity with MMF augments genotoxic chemoradiation therapy in intracranial GBM-bearing mice. (**A**-**C**) Quantification of MPA levels in cortex and tumor tissues harvested from control or MMF-treated mice bearing intracranial GBM38 (**A**), TRP (**B**), or GL26 (**C**) tumors. Data are shown as mean ± SEM of n=5 (**A**), n=6 (**B**), or n=3 (**C**) mice. (**D**) Normalized GTP/IMP ratios in cortex and GBM tissue from mice bearing intracranial GBM38 tumors were determined by LC-MS. Data are shown as mean ± SEM of n=6 biologically independent samples. (**E**-**G**) Intracranial tumors from mice receiving the indicated treatments were stained for γ-H2AX foci and quantified in GBM38 (**E**), TRP (**F**), and GL26 (**G**) models. (**H**-**J**) Quantification of Ki-67 staining in tumors from GBM38 (**H**), TRP (**I**), and GL26 (**J**) tumor-bearing mice receiving the indicated treatments. (**E**-**J**) Data are shown as mean ± SD of n=3-5 biologically independent samples per group. (**K**-**L**) Survival curves of mice bearing intracranial GBM38 (**K**) or intracranial TRP (**L**) tumors receiving the indicated treatments. *p*-values between groups were determined by two-tailed unpaired Student t-tests (**A**-**J**) or log-rank (Mantel-Cox) tests (**K**-**L**). * *p* < 0.05; ** *p* < 0.01; *** *p* < 0.001; **** *p* < 0.0001.

We then sought to understand if MMF could augment the efficacy of RT and TMZ in these GBM mouse models. We first performed mechanistic experiments assessing acute response markers, in which tissues were harvested 4 h after RT, to assess how MMF as monotherapy or in combination with RT/TMZ affected proliferation index and DNA damage repair. MMF treatment alone minimally affected staining of γ-H2AX (Fig. 5E-5G, fig. S6D), a marker of unrepaired DNA, though there was a minor but significant increase in the GBM38 model (Fig. 5E), consistent with the greatest decrease in GTP/IMP ratio in this tumor type. In all three models, combined RT/TMZ expectedly increased γ-H2AX staining in tumors and was further increased with the addition of MMF (Fig. 5E-5G). We observed similar results when assessing staining of Ki-67, a marker of proliferating cells. MMF or RT/TMZ treatment alone decreased Ki-67 tumor staining, while MMF/RT/TMZ combination therapy decreased Ki-67 intensity to the greatest extent, though this combination effect was not significantly different compared to RT/TMZ alone in the GL26 model (Fig. 5H-5J, fig. S6E).

We also conducted therapeutic efficacy experiments to assess how MMF treatment affected tumor growth and survival in mice with intracranial GBM38, TRP or GL26 tumors (fig. S7A-S7C). In the GBM38 model (Fig. 5K), neither RT nor MMF monotherapy meaningfully improved survival. Combined RT/TMZ treatment improved survival compared to RT alone and was similar to combined RT/MMF. The triple combination of RT/TMZ/MMF dramatically improved mouse survival (nearly 7-fold increase compared to untreated mice and 2-fold longer than RT/TMZ alone). In the syngeneic TRP model (Fig. 5L), mice treated with combined RT/MMF and TMZ/RT again had similar survival, which was improved compared to control mice or those treated with RT or MMF monotherapy. Mice treated with the triple combination of RT/TMZ/MMF had the greatest survival, but the increase over standard of care RT/TMZ was more modest than in the GBM38 model. A similar 4-arm experiment in the GL26 model (fig. S7D) showed a trend towards increased survival in MMF/RT/TMZ-treated mice compared to RT/TMZ alone, but this was non-significant. However, monitoring intracranial bioluminescence of GL26 tumors further suggested that combined MMF/RT/TMZ slowed tumor growth compared to RT/TMZ (fig. S7E). In no model did MMF have single agent efficacy. While chemoradiation treatments caused temporary weight loss in tumor-bearing mice (fig. S7F-S7H), MMF treatment did not cause or worsen weight loss when used alone or combined with genotoxic therapy.

### Subhead 6: MMF intracranial exposure in patients with glioma

Finally, we aimed to understand whether the IMPDH inhibitor MMF might be useful for the treatment of patients with GBM. As a first step in evaluation, we performed a phase 0 clinical trial in which patients with recurrent GBM needing a craniotomy for tumor removal received oral MMF for one week prior to surgery with the final dose on the morning of tumor resection (Fig. 6A). At the time of tissue removal, three sample types were obtained using MRI guidance including: (1) contrast-enhancing tumor (typically indicative of aggressive tumor with breakdown of the blood brain barrier); (2) non-enhancing tumor (often indicative of less aggressive tumor with less breakdown of the blood brain barrier); and (3) adjacent cortex tissue, which needed to be removed during the surgery. In addition, a plasma sample was taken at the time of surgery to compare drug levels between tumor and circulation.

**Fig. 6.**
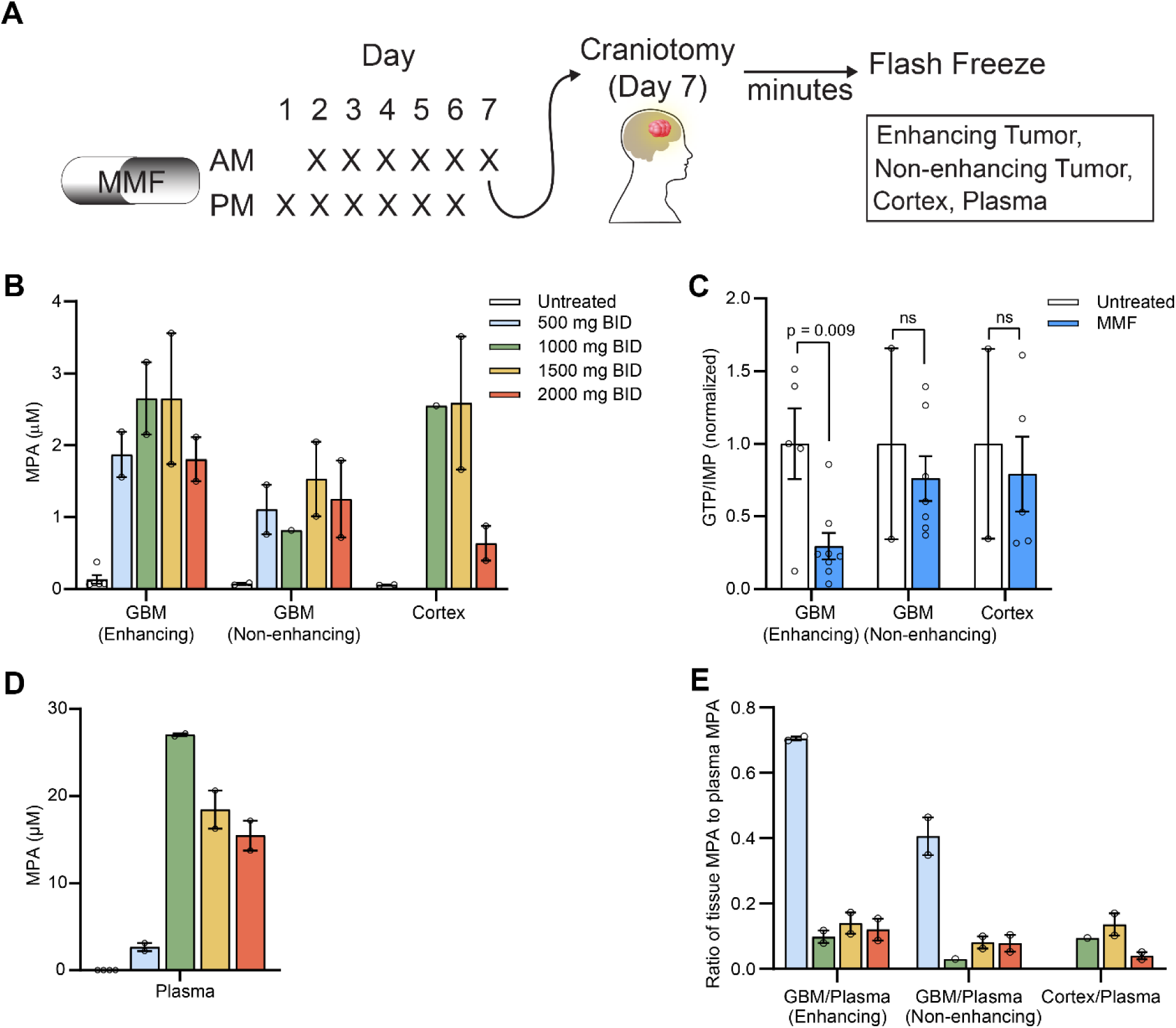
IMPDH can be intracranially targeted with MMF in human patients with GBM. (**A**) Phase 0/1 trial of MMF oral administration and PK/PD analysis in GBM patients: Patients took oral MMF 1-2 times daily for 7 days prior to standard-of-care craniotomy, at which point blood samples and resected tissue samples were collected and analyzed for drug accumulation and target engagement. (**B**) MPA levels in brain tumor (enhancing and non-enhancing tissue) and cortex tissues resected from control patients or patients taking MMF. (**C**) GTP/IMP ratios in brain tumor (enhancing and non-enhancing tissue) and cortex tissues resected from control patients or patients taking MMF. Data are shown as mean ± SEM of n=2-7 biologically independent samples. (**D**) MPA accumulation in plasma samples from control patients or patients taking MMF. Data are presented as mean ± SEM of n=2-4 biologically independent samples per treatment group. (**E**) Ratio of intracranial tissue MPA to plasma MPA in control patients or patients taking MMF. Data are shown as mean ± SEM of n=1-2 biologically independent samples per treatment group. *p*-values between groups were determined by two-tailed unpaired Student t-tests.

Eight patients with recurrent glioblastoma enrolled in this phase 0 study and received MMF 500, 1000, 1500 or 2000 mg BID (n=2 per dose level, Table 1). Patient characteristics including ages, prior treatments, and pathological results are detailed in Table 1. Both patients treated with MMF 2000 mg BID experienced dosing interruptions due to patient error or surgical rescheduling. Five additional samples from contemporaneous patients with recurrent glioblastoma did not enroll in the phase 0 study or receive MMF but underwent tissue harvesting in the same manner and were used as untreated controls.

**Table 1.** Characteristics of patients with glioma studied for pharmacokinetics and pharmacodynamics of MMF.

| Patient | MMF Dose | Sex | Time from initial GBM diagnosis to MMF start | Tissue diagnosis at craniotomy | Extent of resection | Additional information |
| --- | --- | --- | --- | --- | --- | --- |
| 1 | 500 mg BID | F | 3 years, 4 months | OR 1: GBM, MGMT unmethylated<br>OR 2: Gliosarcoma, | OR 1: STR<br>OR 2: STR | Began MMF 7 days prior to OR 2. |
| 2 | 500 mg BID | M | 6 years, 8 months | OR 1: GBM, MGMT unknown<br>OR 2: GBM, with sarcomatous features, MGMT unmethylated | OR 1: STR<br>OR 2: STR | Began MMF 7 days prior to OR 2. |
| 3 | 1000 mg BID | M | 1 year, 6 months | OR 1: GBM, MGMT methylated<br>OR 2: GBM, MGMT methylated | OR 1: GTR<br>OR 2: GTR | Began MMF 7 days prior to OR 2. |
| 4 | 1000 mg BID | F | 9 months | OR 1: GBM, MGMT unmethylated<br>OR 2: GBM, with sarcomatous features | OR 1: GTR<br>OR 2: STR | Began MMF 7 days prior to OR 2. Did not take MMF with RT given low KPS. |
| 5 | 1500 mg BID | F | 7 months | OR 1: GBM, MGMT unmethylated<br>OR 2: Treatment effects | OR 1: GTR<br>OR 2: STR | Began MMF 7 days prior to OR 2. |
| 6 | 1500 mg BID | M | 8 months | OR 1: GBM, MGMT unmethylated<br>OR 2: Treatment effects with minimal residual tumor<br>OR 3: GBM with treatment effects | OR 1: GTR<br>OR 2: GTR<br>OR 3: STR | Began MMF 7 days prior to OR 2. Did not receive MMF prior to OR 3. |
| 7 | 2000 mg BID | M | 1 year, 3 months | OR 1: GBM, MGMT unmethylated<br>OR 2: Gliosarcoma, up to 50% treatment effect | OR 1: GTR<br>OR 2: STR | Began MMF 7 days prior to OR 2; missed one dose. |
| 8 | 2000 mg BID | F | 5 months | OR 1: GBM, MGMT unmethylated<br>OR 2: GBM with treatment effects | OR 1: STR<br>OR 2: GTR | OR 2 delayed 6 days due to physician unavailability. Patient started MMF in initially-scheduled week, stopped, then finished MMF the week of OR 2, as directed. |
| 9 | Control | F | NA | OR 1: GBM, MGMT unmethylated | OR 1: GTR | NA |
|  |  |  |  | OR 2: Recurrent/residual glioma admixed with reactive gliosis and treatment effects | OR 2: STR |  |
|  |  |  |  | OR 3: Gliotic brain tissue with treatment effect | OR 3: STR |  |
| 10 | Control | M | NA | OR 1: Diffuse astrocytoma, IDH mutant, WHO grade II, 1p/19q preserved | OR 1: GTR | NA |
|  |  |  |  | OR 2: IDH mutant, MGMT methylated | OR 2: STR |  |
|  |  |  |  | OR 3: IDH mutant, MGMT methylated | OR 3: GTR |  |
| 11 | Control | M | NA | OR 1: GBM, with focal sarcomatoid differentiation, MGMT unmethylated | OR 1: GTR | NA |
|  |  |  |  | OR 2: Treatment effects with < 10% GBM | OR 2: GTR |  |
| 12 | Control | M | NA | OR 1: GBM, MGMT unmethylated | OR 1: GTR | Sample was resected from patient 6 during a later procedure (OR 3) while not taking MMF. |
|  |  |  |  | OR 2: Treatment effects with minimal residual tumor | OR 2: GTR |  |
|  |  |  |  | OR 3: GBM with treatment effects | OR 3: STR |  |
| 13 | Control | M | NA | OR 1: GBM, MGMT unmethylated | OR 1: STR | NA |
|  |  |  |  | OR 2: GBM, MGMT unmethylated | OR 2: STR |  |

Abbreviations: OR, Operating Room; STR, Subtotal Resection; GTR, Gross Total Resection.

MPA, the biologically active metabolite of MMF, accumulated to approximately 2 μM in contrast-enhancing tumor tissue and to 1-2 µM in the non-enhancing tumor tissue (Fig. 6B). Levels of MPA were similar in the cortex, though we noted modestly lower levels in the cortex tissue from patients treated with 2000 mg BID MMF (both of whom had interruptions in their prescribed MMF dosing). To determine intracranial pathway blockade in humans, we next examined GTP/IMP ratios as in mice to assess for IMPDH inhibition. Enhancing tissue from patients treated with MMF exhibited an approximately 75% decrease in the GTP/IMP ratio compared to untreated patients (p=0.009, Fig. 6C). Receipt of MMF was not associated with a lower GTP/IMP ratio in non-enhancing tumor tissue or in cortical tissue, though wide error bars and limiting sample number (n=2) in control patients likely limited the ability to observe a difference. Absolute levels of the metabolites GTP, GMP, IMP and AICAR did not vary between MMF-treated and control patients, though there was a trend towards decreased GTP (in enhancing tissue) and an increase in the upstream metabolites IMP (in enhancing tissue) and AICAR (in non-enhancing and cortical tissue) in MMF-treated patients (fig. S8A-S8D).

To account for patient biological and treatment variability, we also quantified plasma MPA levels in MMF-treated patients as a normalization factor. Plasma MPA levels ranged between 15 and 30 µM in patients treated with 1000-2000 mg BID MMF and were expectedly lower (2.6 µM) in patients treated with 500 mg BID (Fig. 6D). When intracranial MPA accumulation was normalized to circulating MPA availability, a trend towards increased drug accumulation was observed in enhancing GBM tissue compared to cortex, consistent with observations in tumor-bearing mice (Fig.6E, Fig. 5A-5C). Our lowest studied dose of MMF (500 mg BID) yielded a higher tumor/plasma MPA ratio than higher doses, suggesting that aggressive dose escalation might not yield increased tumor drug exposure. Altogether, these *in vitro*, *in vivo*, and human clinical data reveal a novel mechanism through which GBMs adaptively rewire purine synthesis to resist standard-of-care therapy that can be feasibly targeted in patients with GBM.

## DISCUSSION

In this work, we identify acute changes in purine metabolism following RT in multiple *in vitro* and *in vivo* GBM models suggesting increased purine synthesis. Using stable isotope tracing, we confirm that RT stimulates GBM guanylate synthesis *in vitro* and *in vivo*. These RT-induced metabolic changes depend on an active DNA damage response and are reversed by pharmacologic or genetic inhibition of DNA-PK signaling. Mechanistically, RT-induced DNA-PK activation promotes the nuclear localization of IMPDH1 and an elevation in the GTP/IMP ratio shortly following RT. This biology can be therapeutically targeted. The IMPDH inhibitor MMF accumulates to high concentrations in the cortex and tumor tissue in GBM-bearing mice, where it can decrease GTP/IMP ratio and potentiate the biologic and therapeutic effects of standard-of-care RT and temozolomide. Finally, we performed a phase 0 clinical trial showing that oral MMF dosing leads to active drug concentrations in the tumor tissue of patients with GBM. Together, these studies identify an adaptive metabolic response to RT in GBM that can be interrupted using a well-tolerated FDA-approved oral drug.

These findings have important implications for known links between metabolism and treatment resistance in cancer. Our group and others have previously discovered that GTP promotes resistance to the genotoxic therapies used for glioblastoma treatment (*10–12*). Our observations in this study suggest a compensatory loop where (1) GTP promotes the repair of DNA damage and (2) DNA damage promotes nuclear IMPDH1 localization and GTP synthesis. Similar compensatory loops also exist in breast cancer, where DNA damage promotes the nuclear localization of IMPDH2, which in turn promotes DNA repair (*13*). Though we did not observe nuclear localization of IMPDH2 in our studies following RT-induced DNA damage, it is possible that these different compensatory loops are tissue- or context-dependent. Understanding how GTP is synthesized in different cancer- and treatment-specific situations will likely inform how best to inhibit these metabolic pathways to slow DNA repair and improve the efficacy of genotoxic therapy.

Our experiments in multiple mouse models of GBM indicate substantial heterogeneity in the activity and efficacy of MMF. While MPA, the active metabolite of MMF, accumulated to similar concentrations across models, it had varying effects on metabolite levels and survival. MMF decreased the GTP/IMP ratio (indicating a relative depletion of GTP, which is downstream of IMPDH, and increase of IMP, which is upstream of IMPDH) in the GBM38 model and dramatically improved the efficacy of RT/TMZ. More modest effects on both metabolite levels and survival were observed in the TRP and GL26 models. Together, these studies suggest that the efficacy of MMF in improving survival in GBM models is tied to its ability to alter the GTP/IMP ratio. Because drug penetrance is similar between models, these differences in MMF-induced metabolic changes might be due to different routes of GTP synthesis between models. For example, it is possible that the MMF-sensitive GBM38 model synthesizes the majority of its GTP through IMPDH while the MMF-resistant GL26 model synthesizes more GTP through IMPDH-independent salvage of guanine through HGPRT. Such results could have important implications for patients, as it is possible that those with the greatest IMPDH activity might benefit the most from MMF.

Our phase 0 clinical trial also has implications for the treatment of GBM. There is substantial interest in using IMPDH inhibitors for this disease given several promising preclinical studies of pharmacologic or genetic inhibition of this pathway (*4, 5, 10–12*). While IMPDH inhibitors have been used in patients for decades, their intracranial penetrance is unknown. Our study reveals that oral MMF dosing using standard BID regimens yields relatively high cortical and tumor MPA concentrations. This dosing can lower the GTP/IMP ratio in contrast-enhancing tissue but may not produce similar changes in non-enhancing tumor or normal cortex, which could limit its ability to augment the efficacy of TMZ and RT in non-enhancing tumors. This intracranial penetrance could help explain the efficacy of MMF for the treatment of intracranial inflammatory disorders such as neurosarcoidosis (*39*) and suggests that it might be useful for other diseases such as brain metastases(*18*).

Our study has limitations. While both pharmacologic and genetic approaches to targeting DNA-PK stopped the radiation-induced increase in purine synthesis, we are yet to understand exactly how this signaling pathway promotes the nuclear entry of IMPDH1. Additionally, our phase 0 clinical trial was small and relied on contemporaneous control patients rather than matched measurements pre- and post-treatment on the same patient. As longitudinal tissue analysis becomes more common in GBM, we are optimistic that such studies become possible. This small size of the study made it difficult to assess for MMF-induced changes in non-enhancing tumor and cortical tissue due to the limited number of samples in these regions.

In summary, our study reveals a new link between radiation-induced DNA damage and IMPDH-dependent GTP synthesis. This link can be disrupted by MMF, an orally bioavailable GTP inhibitor, that we now show has excellent CNS penetration in mice and human patients. The heterogeneous benefit of MMF between mouse models suggests that further efforts are needed to directly quantify metabolic fluxes in human cancer to assess which tumors might benefit from inhibition of individual metabolic pathways. These promising preclinical efficacy studies and demonstration of CNS penetrance in humans warrants further testing of IMPDH inhibitors such as MMF in therapeutic clinical trials in brain cancer.

## MATERIALS AND METHODS

### Model sources

DBTRG-05MG (CRL-2020) and A-172 (CRL-1620) cell lines were purchased from ATCC. The GBM38 and GBM06 PDX models were obtained from Dr. Jann Sarkaria (Mayo Clinic) (*19*). The HF2303 primary neurosphere model was originally described by Dr. Tom Mikkelsen at Henry Ford Hospital (*40*). The GL26 model was obtained from Dr. Maria Castro (University of Michigan Medical School) (*36*). The TRP model was obtained from Dr. C. Ryan Miller (The University of Alabama at Birmingham) (*37*).

### Cell culture

Immortalized cell lines and PDX explants were cultured in DMEM (Thermo Fisher Scientific, 11965092) containing 10% FBS (R&D Systems, S11550), penicillin-streptomycin-glutamine (Thermo Fisher Scientific, 10378016), and 100 μg/mL normocin (Invivogen, ant-nr-1). HF2303 neurospheres were cultured in DMEM-F12 (Thermo Fisher Scientific, 10565018) containing B-27 supplement (Thermo Fisher Scientific, 17504044), N-2 supplement (Thermo Fisher Scientific, 17502048), 20 ng/mL EGF (PeproTech, AF-100-15), 20 ng/mL FGF (PeproTech, 100-18B), penicillin-streptomycin (Thermo Fisher Scientific, 15140122), and 100 μg/mL normocin. All cell lines were tested for *mycoplasma* monthly and confirmed negative.

### Animal models

All mouse experiments were approved by the University Committee on Use and Care of Animals at the University of Michigan. B6.129S7-Rag1^tm1Mom^/J mice were obtained from Jackson Laboratory or bred in-house, and C57BL/6J mice were obtained from Jackson Laboratory. Mice were housed in specific pathogen-free conditions at a temperature of 74 °F and relative humidity between 30 and 70% with a light/dark cycle of 12 h on/12 h off. Mice had access to food (PicoLab® Laboratory Rodent Diet 5L0D) and water at all times, except during stable isotope infusions (described below), and were aged 4-12 weeks at the start of intracranial tumor experiments.

Patient-derived and syngeneic tumors were generated by intracranially implanting GFP-positive, luciferase-positive cells into mice with stereotactic guidance (*11*). Human GBM38 and GBM06 tumors were established in B6.129S7-Rag1^tm1Mom^/J mice, and syngeneic mouse-derived TRP and GL26 tumors were established in C57BL/6J mice. Tumor establishment was confirmed via bioluminescence imaging. To measure tumor luminescence, mice were injected with 150 mg/kg D-luciferin intraperitoneally and imaged 10 minutes later with an IVIS Spectrum imaging system (PerkinElmer) under anesthesia (2% isoflurane inhalation). Whole-brain cranial RT was administered to anesthetized (2% isoflurane inhalation) mice that were concealed by a lead shield with only the cranium exposed to the beam. During tissue harvests, intracranial GFP-positive tumor tissue was separated from GFP-negative cortex tissue using guidance by fluorescent light.

### Metabolomic analyses of cells and tissues

Cells or flash-frozen tissues were lysed or homogenized with 80% v/v methanol in water at −80 °C. Plasma samples were mixed with 100% methanol at −80 °C such that final methanol concentration was 80%. Samples were clarified by centrifugation at 4 °C, and supernatants were dried by nitrogen purging or vacuum centrifugation. Dried metabolites were reconstituted with 50% v/v methanol in water for LC-MS. For Snapshot steady-state metabolomics, samples were analyzed using an Agilent QQQ 6470 LC-MS system with multiple reaction monitoring, as described in detail previously (*41*). Data were extracted and analyzed using Agilent MassHunter Quantitative Analysis for QQQ version 10.1. For stable isotope labeling studies of cells and tissues, labeled samples and unlabeled negative controls were analyzed with an Agilent system consisting of an Infinity Lab II UPLC coupled with either a 6545 or 6230 QTOF mass spectrometer (Agilent Technologies, Santa Clara, CA) operated by the University of Michigan Metabolomics Core. Metabolites were identified by matching retention times and mass to authentic standards, with values corrected for natural isotope abundance using Agilent MassHunter Profinder 10.0.

### Pathway Analysis

Pathway enrichment analyses were performed using MetaboAnalyst (https://www.metaboanalyst.ca/, version 6.0) by assessing metabolites with differential abundance between conditions with *p*<0.05. Analysis of intracranial metabolites in the orthotopic HF2303 model were performed with our dataset generated in a parallel study (*20*). The KEGG metabolite library was used for analysis, with backed pathway Holm p (Holm–Bonferroni method correction) for analysis with GraphPad Prism 9.

### Dynamic flux analysis

Dynamic flux analysis (DFA) is based on flux balance analysis (FBA); while FBA assumes steady-state conditions for all metabolites, DFA allows for deviations from steady state for metabolites whose rates of change are measured in time-course metabolomes. Briefly, the time-course metabolomics data from each condition were used as input constraints in the human metabolic reconstruction (*42*) to create condition-specific metabolic models using the DFA approach, as described previously (*21–23*). DFA solves an optimization problem that maximizes the fit to the metabolomics data and an optimization objective, in this case, the biomass, while satisfying the stoichiometric and thermodynamic constraints present in the metabolic network model. DFA outputs a flux solution that best fits all these constraints. DFA models for each condition were compared to identify metabolic reactions that were differentially active between GBM vs brain tissue.

### Stable isotope labeling and treatments *in vitro*

Adherent cells in media containing dialyzed FBS (Thermo Fisher Scientific, A3382001) were labeled with either ^15^N-amide-glutamine (4 mM), 2,8-^2^H_2_-hypoxanthine (100 µM), or 8-^13^C-guanine (100 µM) 30 minutes before RT. Cells were treated with RT (0, 2, or 8 Gy) and then harvested for LC-MS 3 h (GBM38) or 4 h later (A-172). In experiments using DNA damage repair kinase inhibitors, GBM38 cells were pre-incubated with tracer and either M3814 (1 µM), Ku60019 (2 µM), or AZD6738 (0.5 µM) for 3 h before RT (0 or 8 Gy) and then harvested 1 h post-RT for LC-MS or immunoblot. Patient-derived HF2303 neurospheres were incubated with M3814 (1 µM) overnight. For HF2303 neurosphere experiments, following overnight incubation with M3814 (1 µM), media was replaced by fresh media containing tracer and M3814 for 1 h prior to RT (8 Gy), and samples were harvested for LC-MS 2 h post-RT.

### Stable isotope labeling *in vivo*

To establish parameters for stable isotope administration *in vivo* by measuring time-dependent ^15^N-glutamine plasma enrichment, an experiment was performed in which male mice underwent surgical cannulation with one catheter placed into the jugular vein (for tracer infusion) and another placed into the carotid artery (for blood collection). After a recovery period of 4-5 days, conscious, free-moving, undisturbed mice were administered a bolus dose of ^15^N-amide-glutamine (0.28 mg/g) followed by a continuous infusion (5 µg/g/min) via the intravenous line for a total infusion time of 4 h.

During infusions, blood samples were collected via the arterial line into EDTA-coated vials over multiple time intervals for preparation of plasma and then flash-frozen in liquid nitrogen. For studies assessing tissue ^15^N-labeling, an experiment was performed using cannulated female mice with sex-matched intracranial GBM38 tumors. In this experiment, mice were implanted with a third catheter for stomach delivery of vehicle or drug, and the experimental arms comprised vehicle treatment, RT + vehicle treatment, and MMF treatment. Label comparisons between control mice and MMF-treated mice are presented in a separate study (Meghdadi *et al*., *Cell Metabolism*, in press), while labeling in the same control mice vs. RT-treated mice is presented here. Cannulated mice were treated with cranial RT (0 or 8 Gy as described above), removed from anesthesia, and immediately (<5 min post-RT) infused with ^15^N-glutamine as described above. Mice were then sacrificed, and tissues were rapidly harvested on dry ice and flash-frozen in liquid nitrogen for LC-MS analysis.

### Protein extraction and immunoblot

Cells were washed with PBS and lysed on ice using RIPA lysis buffer (89900, Thermofisher) containing PhosSTOP phosphatase inhibitor (04906845001, Roche) and complete protease inhibitor (1187358001, Roche). Protein supernatants were clarified by centrifugation and used to prepare samples for SDS-PAGE. To separate cytosolic and nuclear fractions from cells and tissues, extracts were prepared using NE-PER Nuclear and Cytoplasmic Extraction Reagents (78835, Thermo Fisher) according to the manufacturer’s instructions. Samples of equal protein or tissue weight were subjected to SDS-PAGE followed by electrotransfer to membranes (Millipore) for antibody staining. All antibodies were used at a 1:1000 dilution and obtained as follows: anti-IMPDH1 (35914, Cell Signaling Technology), anti-IMPDH2 (57068, Cell Signaling Technology), anti-β-Actin (sc-47778, Santa Cruz Biotechnology), anti-DNA-PKcs (38168, Cell Signaling Technology), anti-GMPS (sc-376163, Santa Cruz Biotechnology), anti-Caspase-3 (9662, Cell Signaling Technology), anti-Histone H3 (3638S, Cell Signaling Technology), anti-PARP (9542S, Cell Signaling Technology), anti-GART (sc-166379, Santa Cruz Biotechnology), anti-PFAS (61852S, Cell Signaling Technology), anti-ATIC (A304-271A-M, Fortis Life Sciences), anti-APRT (PA5-76741, Invitrogen), anti-HGPRT (PA5-22281, Invitrogen), anti-PPAT (15401-1-AP, Proteintech), and anti-PAICS (A304-546A-M, Fortis Life Sciences).

### Estimations of channeled purine synthesis output

Three to four million cells were seeded per 10-cm culture dish and allowed to grow for 20-24 h in RPMI with 10% dialyzed FBS, after which, X-ray irradiation (8 Gy total dose in Precision X-Ray Irradiator) was performed. To estimate the channeled de novo purine biosynthesis flux by mitochondria-associated purinosomes, ^13^C_3_,^15^N-serine (608130, Millipore Sigma) mediated glycine and formate incorporation into the adenine and guanine nucleotides was estimated as described previously (*30, 34*) (fig. S9A). Briefly, the internalized serine is metabolized in mitochondria, producing ^13^C_2_,^15^N-labeled glycine, and ^13^C-formate. These substrates are preferentially utilized by mitochondria-associated purinosomes, leading to generation of various purine isotopologues—with one glycine and one or two formate mediated label incorporations (fig. S9A). The isotopologue distribution in the nucleotides allows calculation of total channeled purine flux. Post-irradiation, cells were gently washed once with 1x PBS, media was changed to purine-depleted MEM (without serine and glycine, with 10% dFBS) supplemented with ^13^C_3_,^15^N serine to a final concentration of 60 μM, and isotope incorporation was allowed for 4 h. Finally, cells were gently washed thrice with 1× PBS and harvested by scraping after directly adding cold 80% methanol (−20 °C), which causes rapid quenching of enzymatic reactions and extraction of bulk metabolites (*30, 43*). Extracts were dried under N_2_ gas flow, and the dried extracts were reconstituted in 50% acetonitrile with 1 μM chlorpropamide (internal standard) such that the final solution contained extract from ∼1 million cells/10 μL. 2-5 μl of extracts were run on an Orbitrap Fusion Lumos Tribrid Mass Spectrometer, and analyte peak areas were obtained using FreeStyle software (Thermo Fisher Scientific). Each experiment included three to four independent biological replicates.

### Proximity ligation assay (PLA)

PLA was used to determine colocalization of enzyme pairs, in which close spatial proximity of the proteins is assessed using a PLA probe that generates fluorescent puncta when the proteins of interest are <40-60 nm apart (*33*)(fig. S9B). Cells were seeded in DMEM with 10% dialyzed FBS. The next day, cells were treated with RT at dosages of 0 or 8 Gy, replenished with fresh media, and incubated for 4 h before fixation. Cells were fixed with 4% PFA at room temperature for 15min.

PLA was performed as previously described (*44*) following the manufacturer’s protocol. Briefly, fixed cells were rinsed once with PBS and twice with TBS (10 mM Tris, pH 8.0; 150 mM NaCl), and then permeabilized with 0.2% Triton X-100 in TBS for 15 min at room temperature. Following permeabilization, cells were washed with TBS and incubated with PLA blocking solution for 1 h. Primary antibodies against PFAS (Novus, NBP1-84691) and ADSL (Novus, NBP2-03107) were applied for 1 h at room temperature. After removal of primary antibodies, cells were washed twice with PLA wash buffer A. PLA ligation solution was incubated with cells for 30 min at 37 °C in a humidified chamber, followed by two washes with buffer A. Amplification solution was then applied for 2 h at 37 °C. After two washes with PLA wash buffer B, nuclei were stained with DAPI for 5 min. Slides were mounted and prepared for imaging. Nuclei were counterstained with DAPI.

PLA signals were acquired on a Nikon Eclipse TE-2000E inverted microscope (60×/1.49 Apo TIRF objective) with an ORCA-Flash 4.0LT camera. Z-stacks were collected at 130 nm intervals and analyzed using FIJI. Cells with ≥6 PLA puncta were scored positive, and the ratio of positive to total cells (30–50 per condition) was calculated across three independent experiments.

### Pharmacokinetic analyses

Tissue levels of MPA, GMP, GTP, IMP, and AICAR were determined by the University of Michigan Pharmacokinetics Core. Tissues samples were homogenized in 10x PBS and then diluted 4-fold with PBS. For MPA determination, 80 µL of diluted homogenate was mixed with 80 µL acetonitrile and 60 µL internal standard (100 ng/mL MPA-D3 in acetonitrile) for protein precipitation. For determination of GMP, GTP, IMP and AICAR, 80 µL of diluted homogenate was mixed with 120 µL internal standard (100 ng/mL AICAR-^15^N^13^C, GMP-^15^N_5_^13^C_10_, IMP-^15^N_4_, and GTP-D4 prepared in 10% trichloroacetic acid in water) for protein precipitation. Samples were vortexed for 10 min and centrifuged at 3500 rpm for 10 minutes, and supernatants were analyzed by LC-MS/MS. Analytical curves were constructed using 11 nonzero standards by plotting the peak area ratio of metabolites to internal standards against concentration in blank tissue or solvent. Concentrations were calculated via linear regression.

### Immunohistochemistry

Mouse intracranial tissues were fixed in 10% neutral buffer formalin and embedded in paraffin. Samples were sectioned at 4 µm thickness onto slides, which were then deparaffinized in xylene and rehydrated in water, followed by antigen retrieval and blocking. Slides were incubated with primary antibodies anti-Ki-67 (1:5000 dilution; 550609, BD Biosciences) or anti-γ-H2AX (1:1000 dilution; 9718, Cell Signaling Technology) at 4 °C overnight. Slides were then incubated with secondary antibodies for 0.5 hours and stained using the DAB substrate kit (SK-4100, Vector Laboratories). Samples were counterstained with hematoxylin, dehydrated, and mounted. The whole stained sections were scanned with Nikon Microscope Eclipse Ti2 Imaging System. Staining was quantified using ImageJ as described previously (*45*).

### Therapeutic studies *in vivo*

In chemoradiation/MMF combination studies, male and female mice bearing intracranial tumors were treated with MMF (150 mg/kg) and/or TMZ (80 mg/kg) delivered orally 2 h prior to cranial RT (2 Gy/fraction). Whole-brain RT was directed to the cranium by concealing the bodies of anesthetized (2% isoflurane inhalation) mice with a lead shield during treatment. In short-term metabolite and immunohistochemistry experiments, mice were treated as follows: daily MMF treatment for 4 consecutive days, with daily TMZ and RT treatments on the third and fourth days of MMF treatments. Tissues were harvested 4 h after RT on the fourth day.

In therapeutic efficacy experiments, mice were treated as follows: Mice bearing intracranial GBM38 or TRP tumors received treatment with TMZ and MMF once daily for 10 days, with a daily RT fraction administered on days 2-4 and days 7-9 of MMF treatment. Mice bearing intracranial GL26 tumors received one of two different regimens. The first regimen consisted of 9 consecutive days of MMF and TMZ treatment, with RT administered on days 1-3 and 6-8. The second regimen consisted of two treatment cycles: the first cycle was 6 consecutive days of MMF and TMZ (days 1-6) with RT administered on days 2-5, and the second cycle was 3 days of MMF and TMZ on days 9-11 with RT administered on days 9-10.

### Human clinical study

After providing informed consent, twelve patients with diagnosed GBM were enrolled into our IRB-approved clinical study (NCT04477200), in which MMF was taken orally twice each day for 7 days before craniotomies for tumor resection. Plasma samples were drawn on the morning of surgery for MPA quantification using a clinical LC-MS/MS assay. Contrast-enhancing tumor, non-enhancing tumor, and cortex tissue samples were collected during standard-of-care resection, performed by WNA and JH, and then rapidly washed in PBS and immediately flash-frozen in liquid nitrogen by members of the research team for further analysis. GTP/IMP ratios were normalized to the mean value of the untreated control samples measured on the same day. Normalized ratios were compared between treatment groups using a two-sided Student’s t-test assuming equal variances. Patient 6 was included as an MMF-treated patient during his second craniotomy. He later underwent a third craniotomy without MMF tissue removal and did not take MMF.

### Statistical methods

Statistics were performed using R or GraphPad Prism 9 and 10. For metabolite enrichment and comparisons, PLA quantification, γ-H2AX foci formation, Ki-67 staining, groups were analyzed by unpaired two-tailed *t*-tests. Animal survival was determined by the Kaplan-Meier method with groups compared using log-rank (Mantel-Cox) tests.

## Data Availability

All data produced in the present study are available upon reasonable request to the authors.

## Acknowledgments

We are grateful to the patients involved in the study for their generosity to advance medical knowledge. We are also grateful to the neurosurgical team at the University of Michigan for assisting with the collection of clinical samples. We thank Drs. Samuel McBrayer, John Woulfe, Thomas Wubben, and Justin Kollman for valuable discussion. We are also grateful to Drs. Jann Sarkaria, C. Ryan Miller, Maria G. Castro, Alnawaz Rehemtulla, and Ana deCarvalho, and the Henry Ford Health System, for providing patient- and mouse-derived tumor models. We appreciate the technical support provided by Joshua Parsels, Maureen Kachman, Heidi Baum, Thekkelnaycke Rajendiran, Angela Wiggins, and Charles Evans. Animal management, technical and experimental support services were provided by the University of Michigan Unit for Laboratory Animal Medicine (ULAM), the University of Michigan Metabolomics Core (University of Michigan Medical School, Biomedical Research Core Facilities), and the Center for Molecular Imaging at the University of Michigan. VP acknowledges Pennsylvania Department of Health, PennState; and thanks Dr Sergei Koshkin, Metabolomics Core Facility, the Huck Institutes of the Life Sciences. Some illustrations were created using BioRender software.

## Funding

National Institutes of Health grant K99CA300923, F32CA260735, and T32CA009676 (AJS)

National Institutes of Health grant R01GM024129 (VP, SJB)

Rogel Cancer Center Postdoctoral Scholarship (AJS)

Rogel Cancer Center Discovery Award (WZ)

Michigan Radiosensitization SPORE Career Enhancement Program 5-P50-CA-269022-02 (WZ)

NIDDK MMPC-Live grant 1U2CDK135066 (ZW, JF, and NRQ)

CURE Grant (VP)

Huck Innovative and Transformational Seed funds (VP)

---

**Fig. S1.**
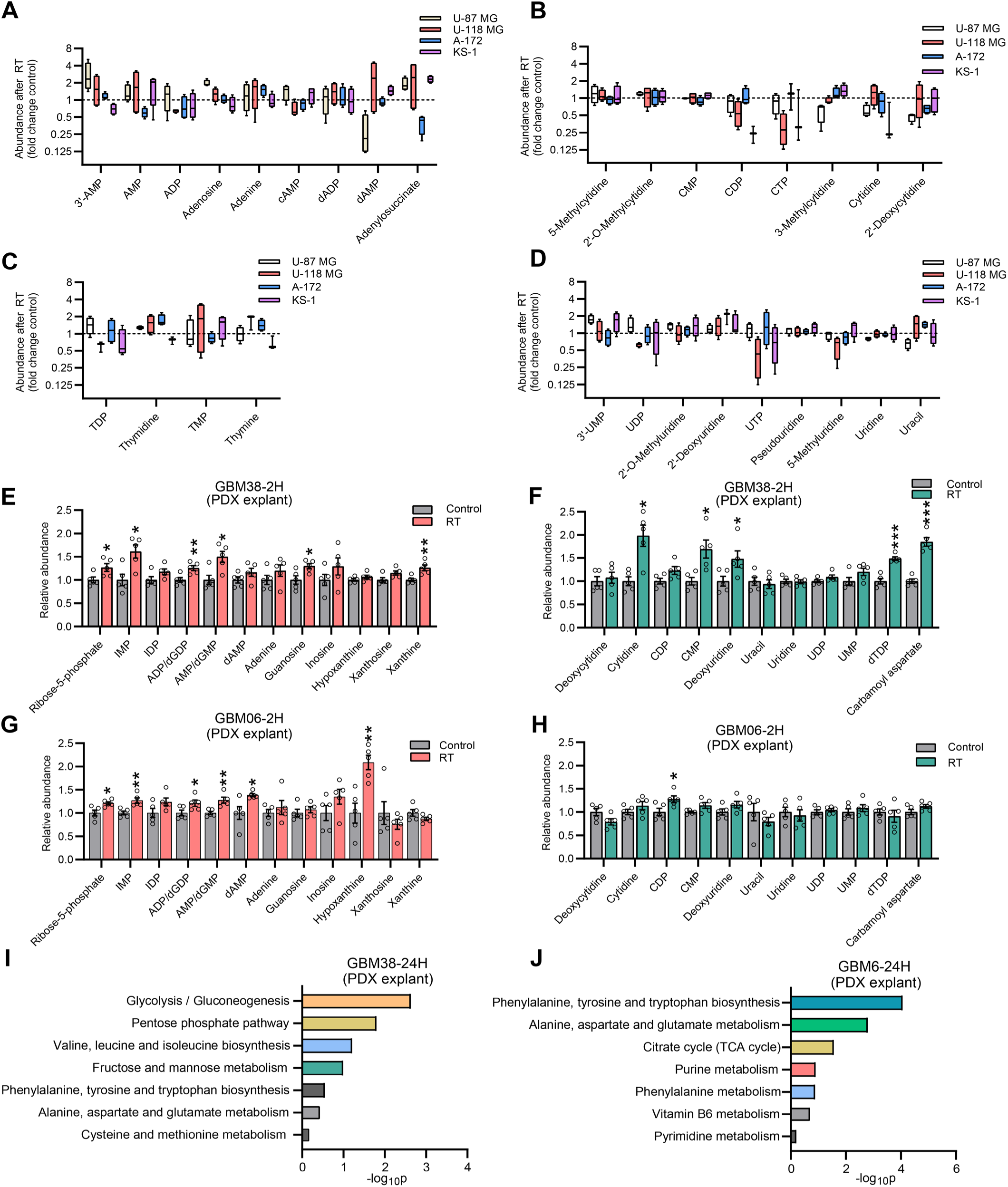
Effect of RT on nucleotide metabolites in GBM models in vitro. (**A**-**D**) Metabolite abundance after RT relative to unirradiated controls: adenylates (**A**), cytidylates (**B**), thymidylates (**C**), and uridylates (**D**). n=4 biologically independent samples per group. (**E**, **F**) Abundance of purine metabolites (**E**) and pyrimidine metabolites (**F**) in GBM38 cells following RT (8 Gy). Data are shown as mean ± SEM of n=5 biologically independent samples per group. (**G**, **H**) Abundance of purine metabolites (**G**) and pyrimidine metabolites (**H**) in GBM06 cells following RT (8 Gy). Data are presented as mean ± SEM of n=5 biologically independent samples per group. (**I**, **J**) Metabolite pathway enrichment analysis of differential metabolites 24 h after RT (8 Gy) in GBM38 (**I**) and GBM06 cells (**J**). *p*-values between groups were determined by two-tailed unpaired Student t-tests. * *p* < 0.05; ** *p* < 0.01; *** *p* < 0.001.

**Fig. S2.**
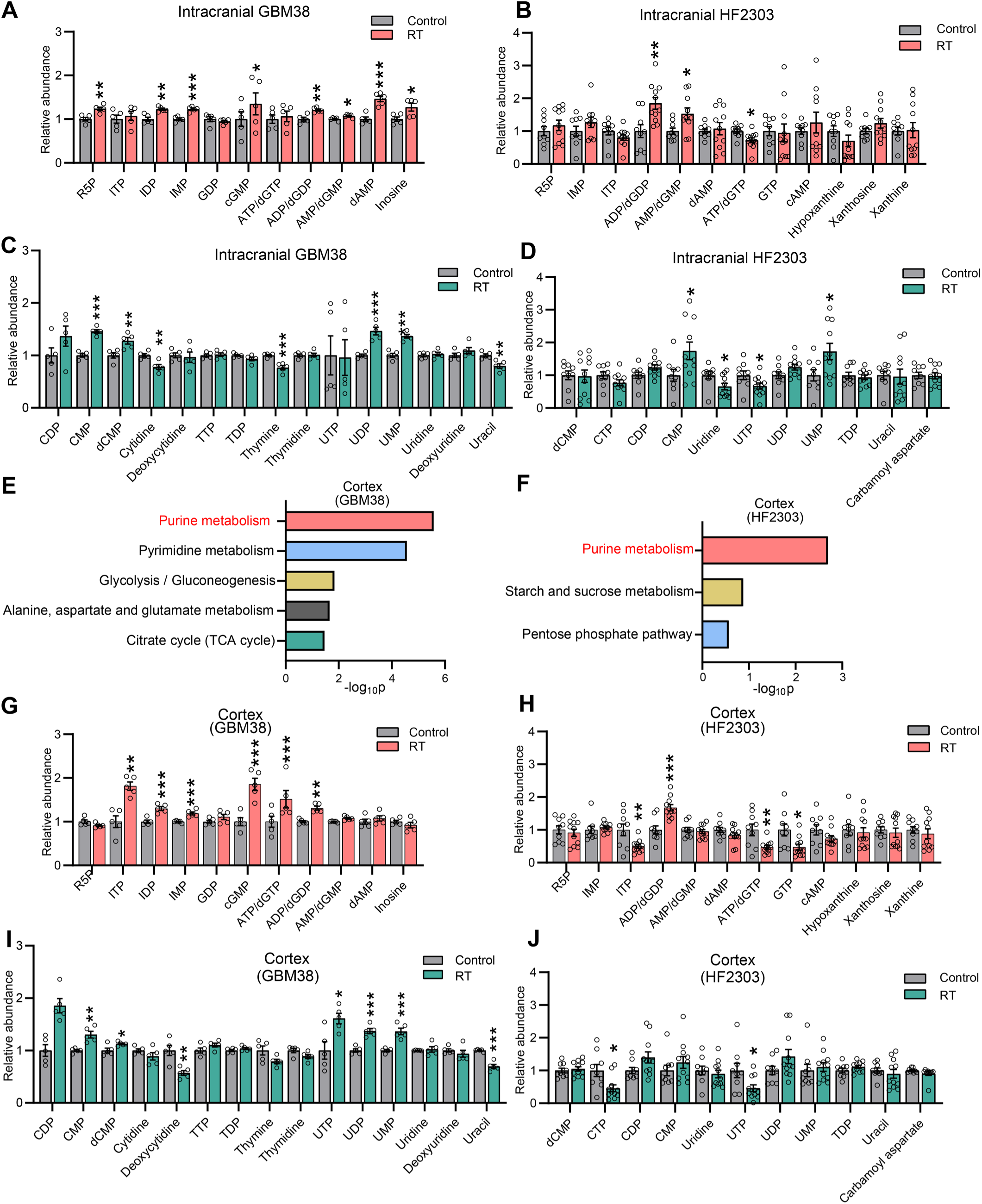
Effect of RT on intracranial nucleotides in orthotopic GBM models. (**A**, **B**) Abundance of purine metabolites in orthotopic GBM38 tumors (corresponding to main Fig. 1E) from mice treated with cranial RT (8 Gy) that were harvested 2 h later (**A**), and in orthotopic HF2303 tumors (corresponding to Fig 1F) from mice treated with RT (2 Gy x 2) that were harvested 4 h later (**B**). Data are shown as mean ± SEM of n=5 (**A**) or n=11 (**B**) biologically independent samples per group. (**C**, **D**) Abundance of pyrimidine metabolites in orthotopic GBM38 tumors (**C**) and HF2303 tumors (**D**). Data are shown as mean ± SEM of n=5 biologically independent samples per group (**C**) or n=11 (**D**) for (**E**-**F**) Pathway enrichment analysis of differential metabolites after RT in cortex tissues from orthotopic GBM38-bearing mice (**E**) and orthotopic HF2303-bearing mice (**F**). (**G**, **H**) Abundance of purine metabolites in cortex tissues from orthotopic GBM38-bearing mice (**G**) and cortex tissues orthotopic HF2303-bearing mice (**H**). Data are shown as mean ± SEM of n=5 (**G**) or n=11 (**H**) biologically independent samples per group. (**I**, **J**) Abundance of pyrimidine metabolites in cortex tissues from orthotopic GBM38-bearing mice (**I**) and cortex tissues from orthotopic HF2303-bearing mice (**J**). Data are shown as mean ± SEM of n=5 (**I**) and n=11 (**J**) biologically independent samples. *p*-values between groups were determined by two-tailed unpaired Student t-tests. * *p* < 0.05; ** *p* < 0.01; *** *p* < 0.001.

**Fig. S3.**
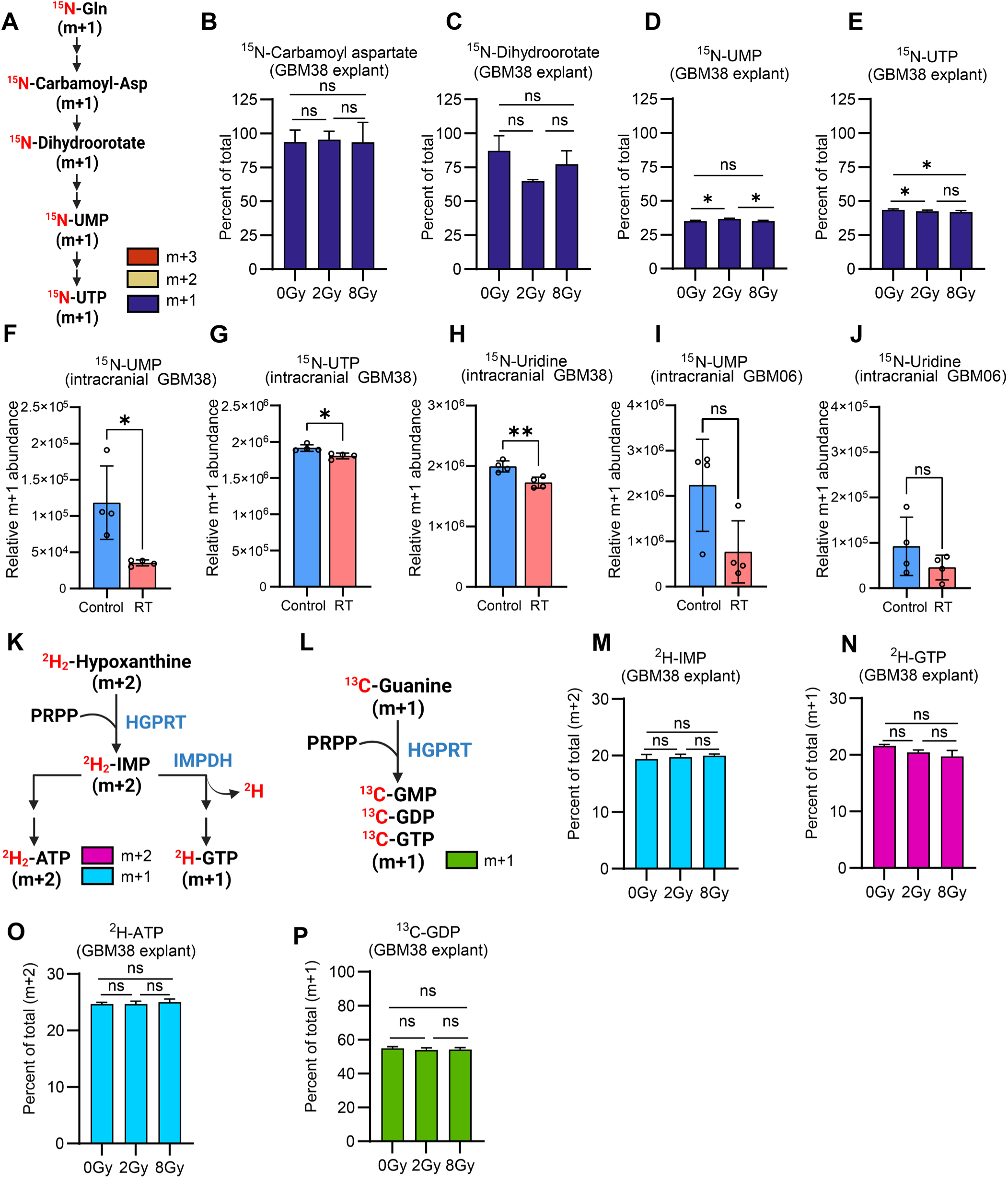
Effect of RT on GBM nucleotide synthesis pathway activity. (**A**) Nitrogen labeling patterns of *de novo* pyrimidine intermediates from ^15^N-amide-glutamine. (**B**-**E**) Abundance of m+1 isotopologue of carbamoyl aspartate (**B**), dihydroorotate (**C**), UMP (**D**) and UTP (**E**) in ^15^N-labeled GBM38 cells treated with RT (0, 2, or 8 Gy). Data are shown as mean ± SD from 4-5 biologically independent samples per group. (**F**-**H**) Abundance of m+1 isotopologue of UMP (**F**), UTP (**G**) and uridine (**H**) in orthotopic GBM38 tumors harvested from mice that were treated with cranial RT (0 or 8 Gy) and immediately infused with ^15^N-glutamine for 4 h. Data are shown as mean ± SD of n=4 biological replicates per group. (**I**-**J**) Abundance of m+1 isotopologue of UMP (**I**) and uridine (**J**) in orthotopic GBM06 tumors harvested from mice that were treated with cranial RT (0 or 8 Gy) and immediately infused with ^15^N-glutamine for 4 h. Data are shown as mean ± SD of n=4 biological replicates per group. (**K**) Labeling of salvaged purines by 2,8-^2^H_2_-hypoxathine: both deuterium atoms are retained during condensation of hypoxanthine and PRPP into IMP. Downstream adenylates retain both deuterium atoms, while during guanylate synthesis one deuterium atom is released in the IMPDH reaction. (**L**) Labeling of salvaged guanylates by 8-^13^C-guanine. Mass isotopologue distributions of IMP (**M**), GTP (**N**), and ATP (**O**) in 2,8-^2^H_2_-hypoxathine-labeled GBM38 cells treated with RT (0, 2, or 8 Gy). Data are shown as mean ± SD of 5 biologically independent samples. (**P**) Abundance of m+1 isotopologue of GDP in 8-^13^C-guanine-labeled GBM38 cells treated with RT (0, 2, or 8 Gy). Data are shown as mean ± SD of 3 biologically independent samples. *p*-values between groups were determined by two-tailed unpaired Student t-tests. * *p* < 0.05; ** *p* < 0.01; *** *p* < 0.001.

**Fig. S4.**
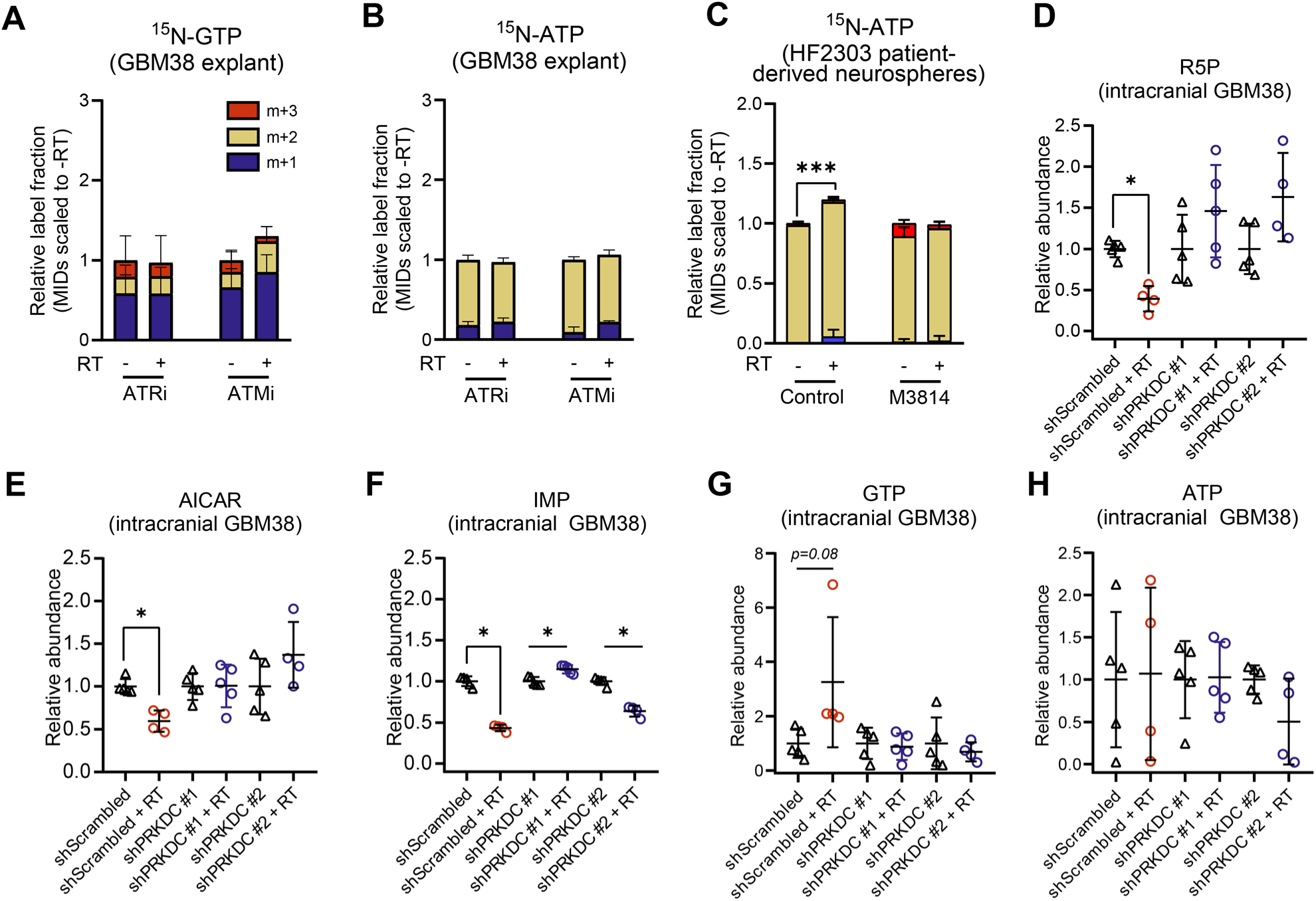
Effects of blocking DNA damage signaling on RT-induced purine metabolism. (**A**, **B**) Mass isotopologue distributions of GTP (**A**) and ATP (**B**) in ^15^N-glutamine-labeled GBM38 cells treated with RT (0 or 8 Gy) and AZD6738 (ATRi, 0 or 50 nM) or KU60019 (ATMi, 0 or 2 µM ATMi). Data are shown as mean ± SD from 3-4 biologically independent samples per group. (**C**) Mass isotopologue distribution of ATP in ^15^N-glutamine-labeled HF2303 neurospheres treated with RT (0 or 8 Gy) and M3814 (0 or 1 µM). Data are shown as mean ± SD from 3-4 biologically independent samples per group. (**D**-**H**) Abundance of R5P (**D**), AICAR (**E**), IMP (**F**), GTP (**G**), and ATP (**H**) in shRNA-modified orthotopic GBM38 tumors from mice treated with cranial RT (0 or 8 Gy). Data are shown as mean ± SD of n=4-5 biologically independent samples per group. *p*-values between groups were determined by two-tailed unpaired Student t-tests. * *p* < 0.05; ** *p* < 0.01; *** *p* < 0.001.

**Fig. S5.**
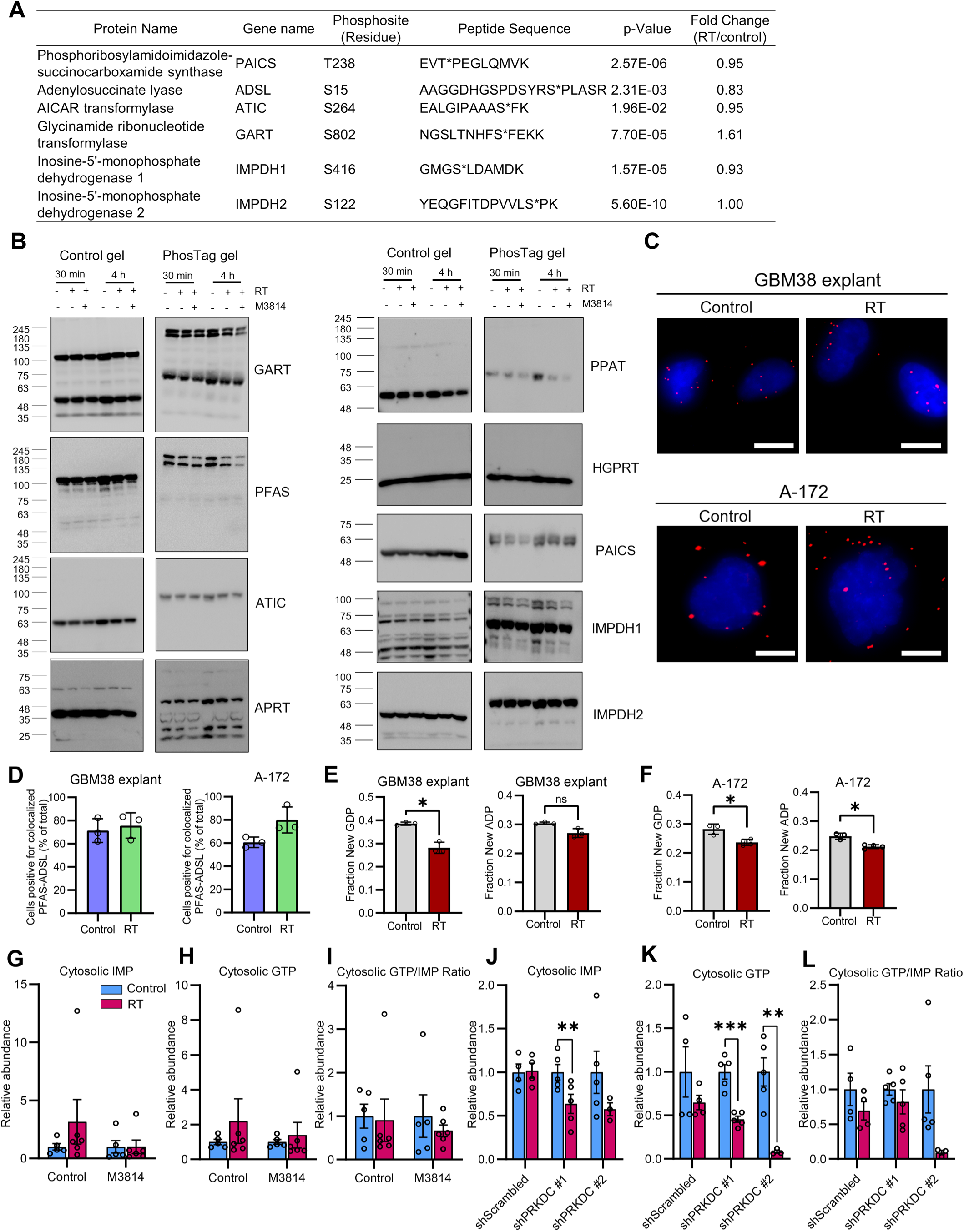
Impact of RT and DNA-PK inhibition on enzyme phosphorylation and subcellular metabolism. (**A**) Phosphorylation of purine biosynthesis enzymes post-RT were determined from a previous phosphoproteomic experiment (*1*). (**B**) Immunoblots for purine synthesis enzymes with phosphorylation-dependent gel shifts were performed to assess purine biosynthesis enzymes in GBM38 cells subjected to indicated conditions. (**C**) PLA was performed to determine colocalization of PSAT and ADSL 4 h post-RT (0 or 8 Gy) in GBM38 explant cells and A-172 cell lines. Red puncta represent colocalized enzyme pairs with blue DAPI counterstaining. Scale bar represents 10 µm. (**D**) Percentage ratios of puncta-positive (≥6 puncta) cells to total cells (30-50 per condition) were calculated across three independent PLA experiments assessing GBM38 explants and A-172 cells. (**E**-**F**) Estimations of GDP and ADP fractions synthesized via purinosomal channeling were calculated using mass isotopologue distributions of metabolites from cells labeled with ^13^C_3_,^15^N-serine that were harvested 4 h post-RT (0 or 8 Gy). Data are shown as mean ± SD of n=3-4 biologically independent samples. (**G**-**I**) Cytosolic fractions of GBM38 cells treated with M3814 or RT (corresponding to Fig. 4E-G) were assessed for IMP (**G**), GTP (**H**), and GTP/IMP ratios (**I**) by LC-MS. Values were normalized to 0 Gy-treated vehicle/drug-only control groups. Data are shown as mean ± SEM of n=5-6 biologically independent samples. (**J**-**L**) Cytosolic fractions (corresponding to Fig. 4H-J) of intracranial GBM38-shScrambled or GBM38-shPRKDC tumors treated with RT (0 or 8 Gy) were assessed for IMP (**J**) and GTP (**K**) by LC-MS and used to determine GTP/IMP ratios (**L**). Data were normalized to 0 Gy-treated shScrambled tumors. Data are shown as mean ± SEM of n=3-5 biologically independent samples. *p*-values between groups were determined by two-tailed unpaired Student t-tests. * *p* < 0.05; ** *p* < 0.01; *** *p* < 0.001.

**Fig. S6.**
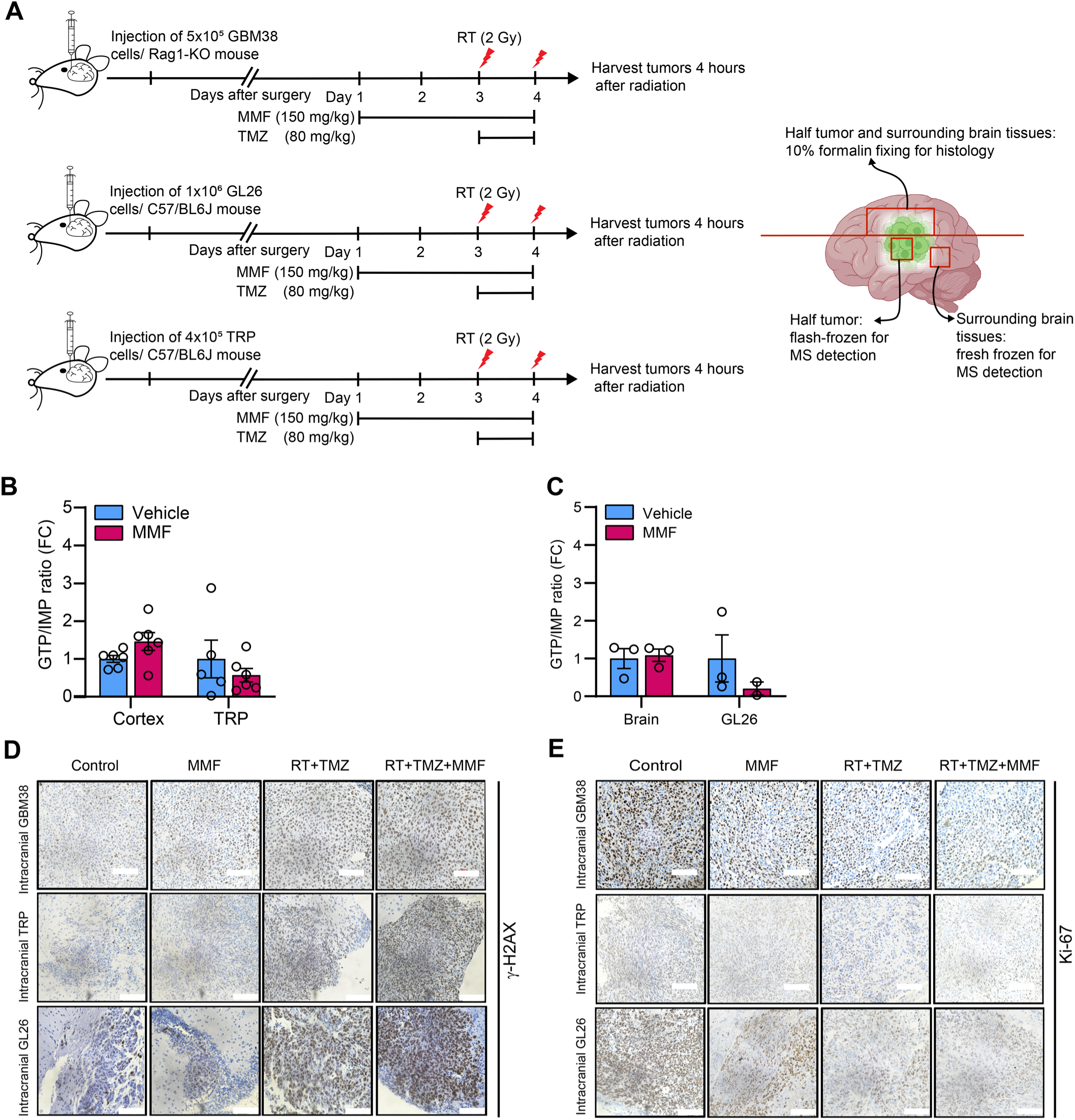
Intracranial targeting of IMPDH with MMF in combination with chemoradiation in GBM-bearing mice. (**A**) Schematic of treatment timelines and tissue harvests for GBM38, TRP, GL26 intracranial mouse models. (**B**, **C**) Normalized GTP/IMP ratios in cortex and GBM tissue from mice bearing intracranial TRP (**B**) or GL26 (**C**) tumors were determined by LC-MS. Data are shown as mean ± SEM of n=6 (**B**) or n= 2-3 (**C**) biologically independent samples. (**D**, **E**) Representative immunohistochemical γ-H2AX (**D**) and Ki-67 (**E**) stains of tumor tissue from intracranial GBM38, TRP, and GL26 tumor-bearing mice (corresponding to Fig. 5E-5J). Scale bar: 100 μm.

**Fig. S7.**
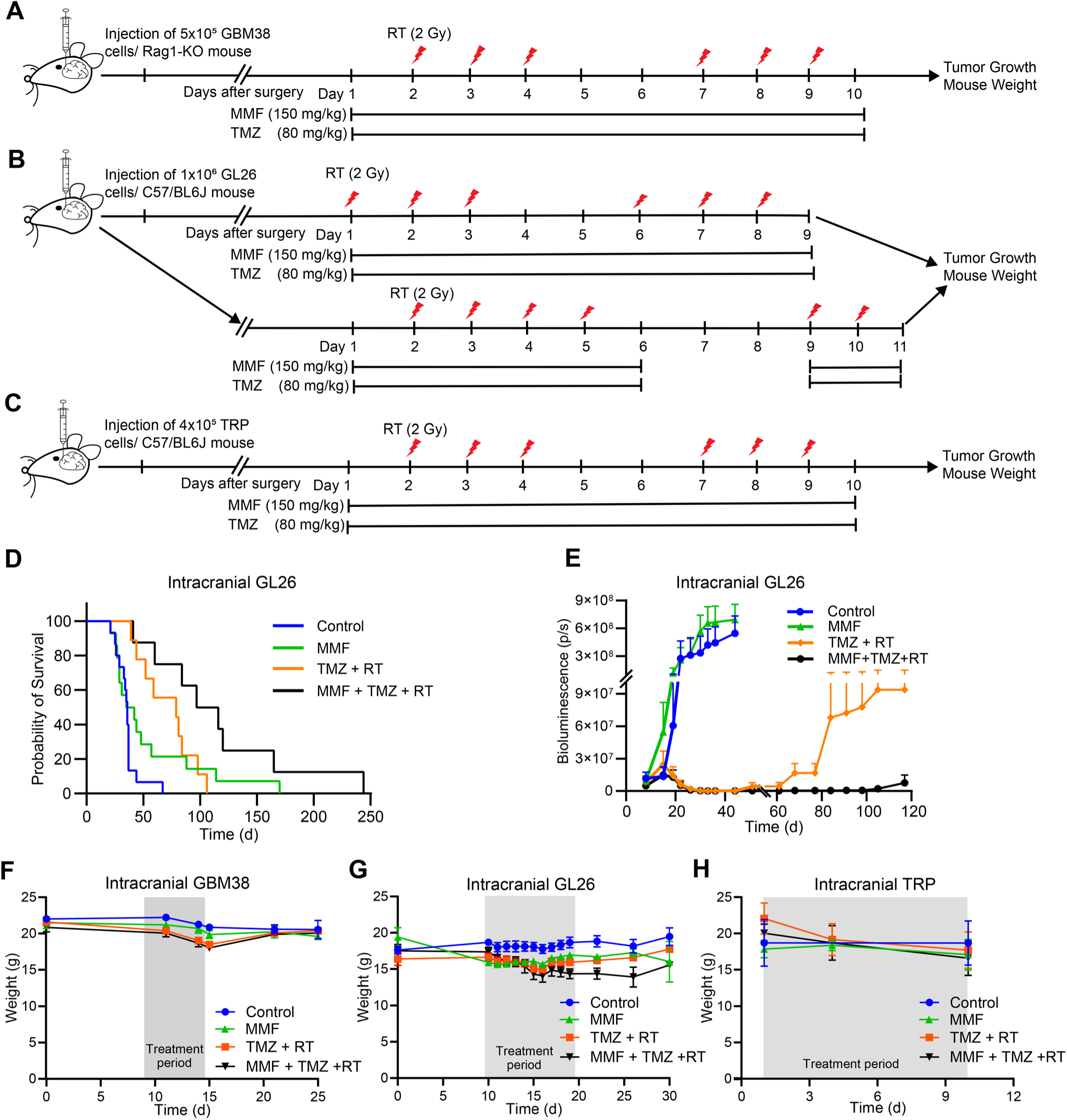
Analysis of combined MMF and chemoradiation treatments in brain tumor-bearing mice. (**A**-**C**) Treatment timelines for GBM38 (**A**), TRP (**B**), GL26 (**C**) intracranial mouse models. (**D**) Survival of intracranial GL26-tumor bearing mice receiving the indicated treatments. (**E**) Bioluminescence fluxes of equal-area regions around crania of intracranial GL26-bearing mice receiving indicated treatments. (**F**-**H**) Bodyweights of orthotopic GBM38-bearing mice (**F**), intracranial GL26-bearing mice (**G**) and TRP-bearing mice (**H**) receiving the indicated treatments. Data are shown as mean ± SEM of n=3-6 per group.

**Fig. S8.**
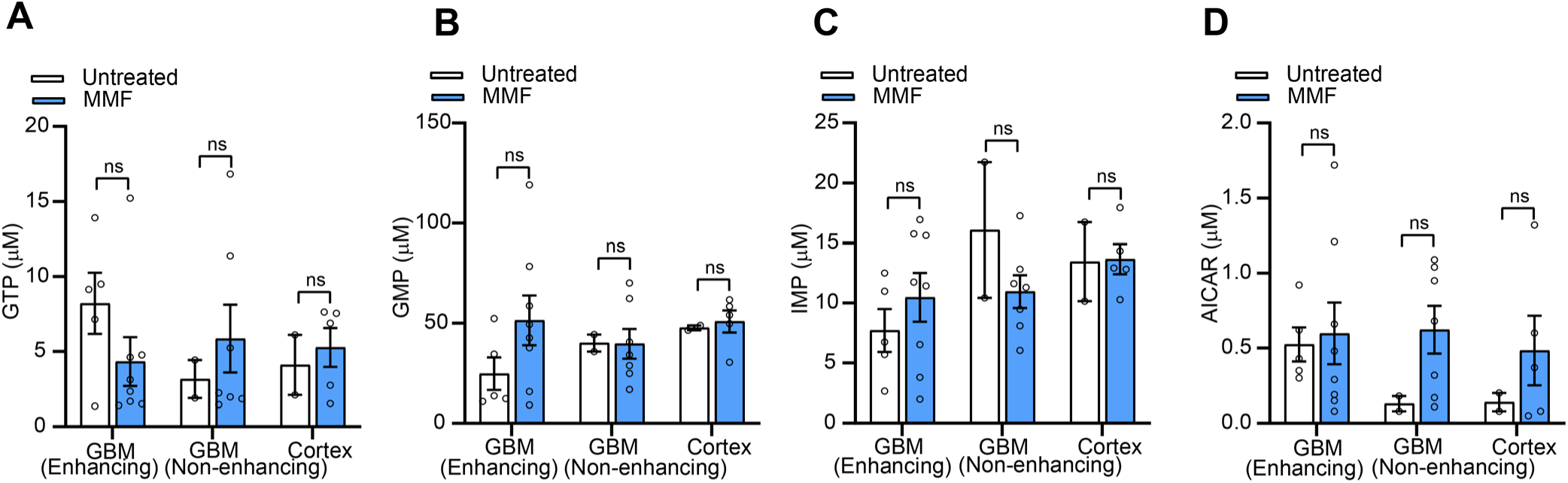
Effect of MMF on intracranial tissue metabolites in human patients. Concentrations of GTP (**A**), GMP (**B**), IMP (**C**), and AICAR (**D**) in intracranial tissues resected from patients taking MMF or non-trial patients. Data are shown as mean ± SEM of n=2-8 biologically independent samples. Comparisons between groups were performed by two-tailed unpaired Student t-tests.

**Fig. S9.**
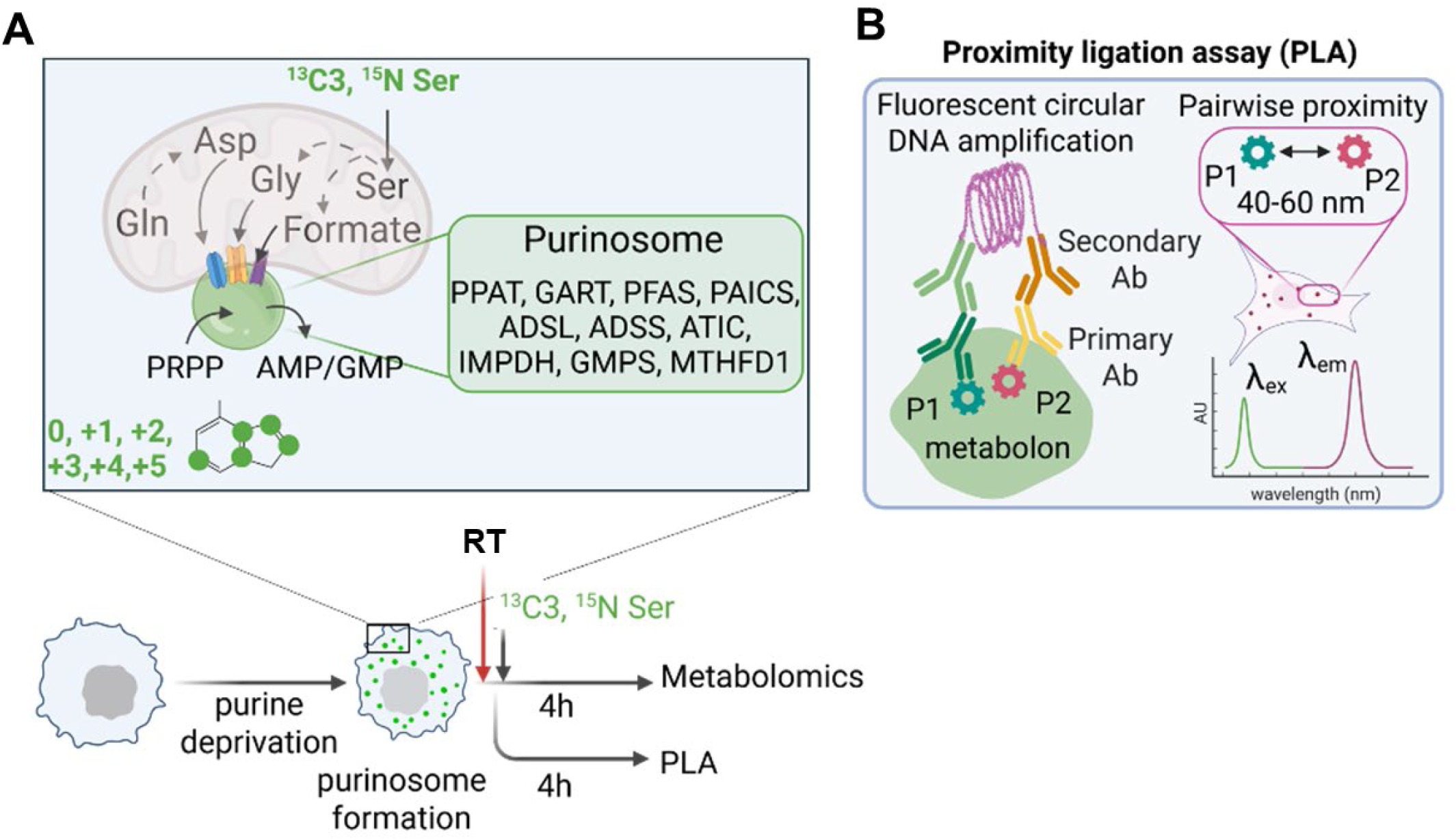
Assessment of purinosome-mediated purine biosynthesis. (**A**) Schematic depicting RT treatment of cells, followed by metabolic assessment or proximity ligation assay (PLA). After growth in purine-depleted media to induce purinosome formation, cells were treated with RT (0 or 8 Gy), and purinosome formation and function were assessed. ^13^C_3_,^15^N-Serine-mediated labeling of purines generated by purinosome-mediated de novo purine biosynthesis was estimated by quantitative metabolomics. Enzymes constituting the purinosomes and the substrates derived from mitochondrial metabolism are shown in the inset. (**B**) Schematic describing PLA, a fluorescence imaging-based assay to assess the proximity of a pair of proteins of interest (P1= PFAS; and P2= ADSL) involved in the purinosome assembly. When the probed pair of proteins is within 40-60 nm distance, it leads proximity of their respective primary and modified secondary antibodies, allowing fluorescent circular DNA amplification by polymerase chain reaction. Therefore, appearance of fluorescent spots reports on the proximity of proteins involved in purinosome formation.

